# Hiporfin (HpD) for fluorescence-guided resection of recurrent glioblastoma: diagnostic accuracy and feasibility of an integrated photodynamic strategy

**DOI:** 10.64898/2026.10.07.26364687

**Authors:** Zhenqiang He, Guanhua Zhang, Changyu Wang, Luyao Wu, Wanming Hu, Hao Duan, Yinsheng Chen, Jie Lu, Ze Yuan, Xiangheng Zhang, Yonggao Mou

## Abstract

**Background:** Hematoporphyrin derivative (HpD, Hiporfin) is used for photodynamic therapy (PDT) of solid tumors but had never been evaluated for fluorescence-guided resection (FGR) in glioma. We assessed Hiporfin fluorescence accuracy against histopathology in recurrent glioblastoma (rGBM) and the feasibility of adding cavity PDT to FGR.

**Methods:** In this prospective single-arm study, adults with rGBM received a single preoperative intravenous Hiporfin dose of 5 mg/kg before FGR under blue-light excitation and 630-nm cavity PDT. Intraoperative fluorescence assessment was the index test and histopathology the reference standard. Specimen-level diagnostic accuracy was pooled with bivariate random-effects modeling.

**Results:** Of 63 specimens, 35 were true positive, 7 false positive, 8 false negative and 13 true negative. Pooled sensitivity was 0.751 (95% confidence interval 0.600–0.858) and specificity 0.606 (0.407–0.775); the crude positive predictive value was 0.833 (0.694–0.917) and the negative predictive value 0.619 (0.409–0.792). All seven false-positive specimens were peritumoral tissue and the eight false negatives were non-fluorescent tumor. FGR combined with cavity PDT was completed in all six patients without new neurological deficits.

**Conclusions:** Hiporfin fluorescence identified rGBM with moderate accuracy: fluorescent regions were likely tumor, while non-fluorescent tissue often still contained tumor. The main source of error was peritumoral tissue of the treated cavity, and not the drug or the optics.

Hiporfin is a new fluorescence agent for glioma. One drug provides both fluorescence guidance during resection and immediate cavity PDT.

**Key Points:**

- Hiporfin fluorescence in recurrent glioblastoma: sensitivity 0.75, specificity 0.61.
- Fluorescence predicted tumor with PPV 0.83, but its absence had NPV 0.62.
- First specimen-level accuracy estimated for Hiporfin fluorescence in recurrent glioblastoma.

**Importance of the Study:** Hiporfin (hematoporphyrin derivative) is used for photodynamic therapy of solid tumors, but its fluorescence had not been characterized in the operative field. We provide the first clinical fluorescence-guided resection data for this agent in glioma and pair it with immediate cavity photodynamic therapy in a single procedure. In this prospective six-patient series, pooled sensitivity was 0.75 and specificity 0.61, so more than a third of non-fluorescent specimens still contained tumor while every false-positive specimen was peritumoral tissue. That failure mode is intrinsic to reoperation in a treated cavity and not to the drug or the optics. The workflow was completed in every patient without new deficit; the one patient with both a newly diagnosed and a recurrent lesion suggests greater potential for Hiporfin in newly diagnosed glioblastoma, which warrants prospective evaluation.

## Introduction

Almost all patients with glioblastoma experience recurrence despite surgery, radiotherapy, chemotherapy and even tumor-treating fields^1,2^. Standards of care at recurrence are less well defined than at diagnosis. Both the EANO and the NCCN guidelines individualize salvage treatment according to prior therapy, age, Karnofsky performance status (KPS), O⁶-methylguanine-DNA methyltransferase (MGMT) promoter methylation and progression pattern^3,4^. Neither recommends any single treatment, and both encourage consideration of a clinical trial. A meta-analysis of more than 10,000 recurrent glioblastoma (rGBM) patients found that complete resection of the enhancing tumor was associated with longer post-recurrence survival^5^. Repeat resection is therefore offered to selected patients. At repeat resection, the difficulty is that residual tumor, radiation necrosis and gliotic scar are difficult to distinguish under white light, so the extent of resection rests on the surgeon’s judgement. Fluorescence-guided resection (FGR) distinguishes glioma from the surrounding tissue, and photodynamic therapy (PDT) to the cavity immediately afterwards may improve local control.

Fluorescence-guided resection makes tumor visible in the operative field, helping the surgeon judge tumor margins and achieve complete resection. 5-aminolevulinic acid (5-ALA) is the established agent: in the randomized trial, complete resection of contrast-enhancing tumor was achieved in 65% of patients given 5-ALA versus 36% with white light, and 6-month progression-free survival was 41.0% versus 21.1%^6^. Its limitations are real. Protoporphyrin IX accumulates in reactive and inflammatory tissue, and 5-ALA labeling is not restricted to tumor cells^7-9^. In patients with newly diagnosed glioblastoma, the positive predictive value of fluorescence was 100% for strong fluorescence and 92.2% for weak fluorescence in pooled biopsies^10^. And all seven false positives were weakly fluorescent. Where tumor cells are absent, fluorescence-positive specimens are more frequent in recurrent than in newly diagnosed glioblastoma^11^.

FGR with 5-aminolevulinic acid has been combined with intraoperative PDT in recurrent glioblastoma in a small case series and a matched cohort study^12,13^. Whether this strategy transfers to other photosensitizers is unknown. Porphyrins are simultaneously diagnostic and therapeutic. Hematoporphyrin derivative (HpD), known as Hiporfin, has been used for PDT of solid tumors^14,15^. The same molecule emits red fluorescence under blue-violet excitation and generates cytotoxic reactive oxygen species under red illumination^16,17^. Although Hiporfin based PDT in glioma was reported previously, the agent has never been used for FGR in glioma^18^.

We therefore undertook a prospective single-arm feasibility study of an integrated strategy in recurrent glioblastoma. The strategy combined a single preoperative 5 mg/kg dose of Hiporfin, fluorescence-guided resection under blue-light excitation, and immediate 630 nm illumination of the resection cavity in the same operation. The aim of this study was to quantify the diagnostic performance of Hiporfin fluorescence at the specimen level against histopathology, which has not been reported for this agent. For established agents, quantitative fluorescence has been related to proliferation markers or cell density, and not to whether a specimen contains tumor^19,20^. We also investigated whether this one-drug, one-dose, one-wavelength FGR plus PDT workflow is feasible with a different photosensitizer, which would make Hiporfin a new option for the strategy. The aim of this study was to quantify the diagnostic performance of Hiporfin fluorescence, used as an adjunct for intraoperative margin guidance during repeat resection, at the specimen level against histopathology, which has not been reported for this agent.

## Methods

### Study design

This prospective, single-arm study was performed at a single cancer center and includes all patients in our institutional rGBM program who received FGR with Hiporfin followed by intraoperative PDT in the same procedure (Figure 1A and 1B). The primary endpoint was diagnostic accuracy, with descriptive early-feasibility secondary endpoints, and the study is reported in accordance with STARD 2015^21^. Efficacy outcomes were specified in the parent study but are not reported here. The study was conducted with institutional ethics approval (No. B2021-199-01) and registered at ClinicalTrials.gov (NCT07832448). Written informed consents were obtained from all patients, including for the research use of de-identified operative video and pathology material.

**Figure 1.**
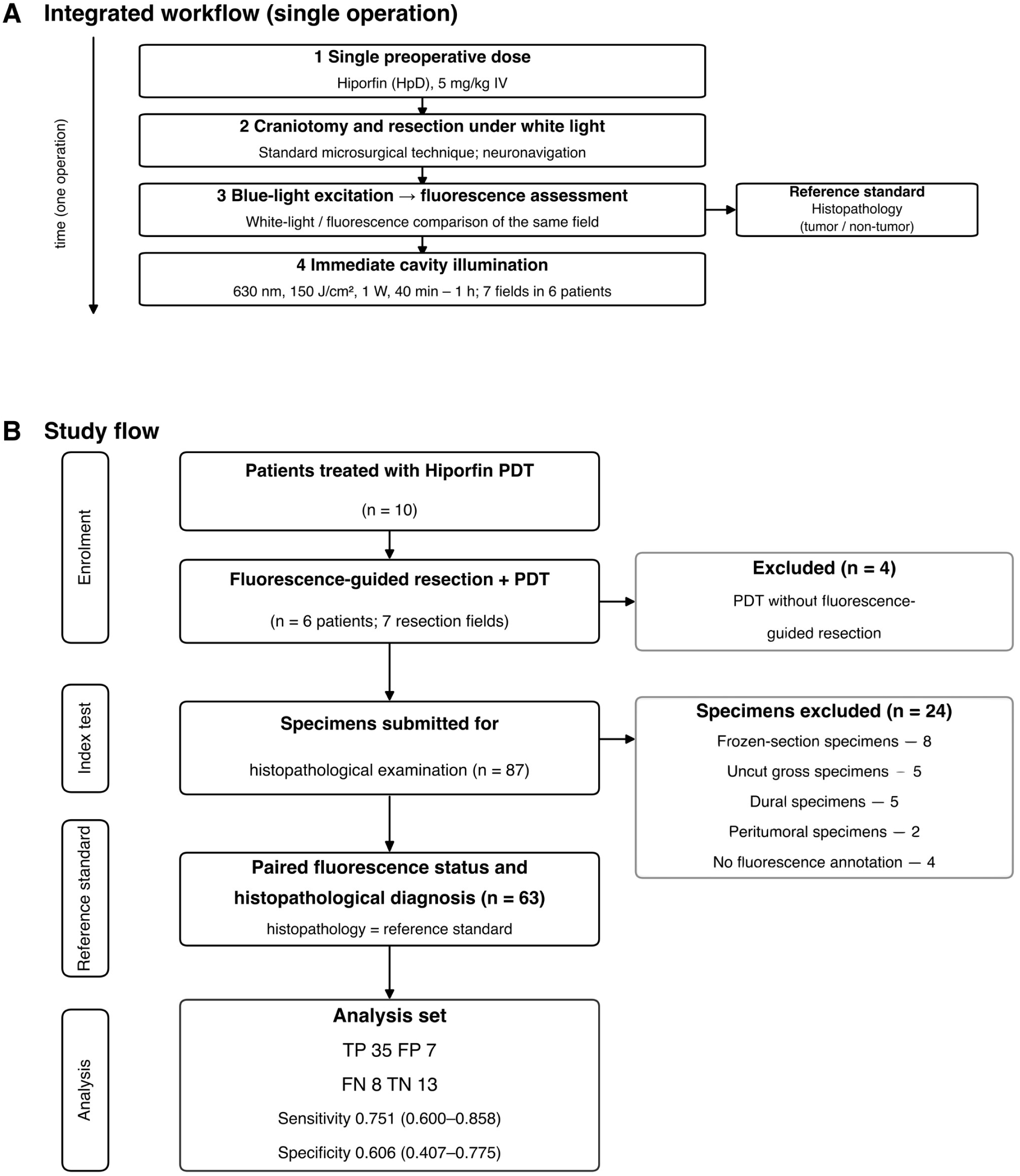
Integrated workflow and study flow. **A**, Fluorescence navigation and photodynamic therapy delivered from a single preoperative dose of Hiporfin (hematoporphyrin derivative, 5 mg/kg intravenously): craniotomy and resection under white light with neuronavigation, blue-light excitation with fluorescence assessment of the same operative field, and immediate cavity illumination (630 nm, 150 J/cm², 1 W, 40 min–1 h; seven illuminated fields in six patients). Histopathology served as the reference standard for every specimen. **B**, Study flow (STARD diagram). Ten patients treated with Hiporfin photodynamic therapy were screened and four were excluded because they received photodynamic therapy without fluorescence-guided resection. Six patients (seven resection fields) underwent fluorescence-guided resection with paired specimen sampling, yielding 87 specimens submitted for histopathological examination. Twenty-four specimens were excluded: frozen-section specimens (8), uncut gross specimens (5), dural specimens (5), peritumoral specimens (2) and specimens without an intraoperative fluorescence annotation (4), leaving 63 specimens with a paired intraoperative fluorescence status and histopathological diagnosis. The analysis set comprised 35 true-positive, 7 false-positive, 8 false-negative and 13 true-negative specimens; pooled sensitivity was 0.751 (95% CI 0.600–0.858) and pooled specificity 0.606 (95% CI 0.407–0.775). HpD = hematoporphyrin derivative; PDT = photodynamic therapy; STARD = Standards for Reporting of Diagnostic Accuracy.

### Patients

Potentially eligible patients were identified prospectively. Those with previously histopathologically confirmed glioblastoma who presented with brain MRI findings suggestive of local recurrence were referred to the study team for screening. Patients in the parent study who received PDT without FGR were not eligible for this analysis. Every anatomically distinct lesion resected was counted as one resection field.

Main inclusion criteria were prespecified in the registered protocol: (1) Age 18 to 65 years. (2) Histopathologically confirmed glioblastoma recurring or progressing after standard radiotherapy and chemotherapy^22^. (3) A Karnofsky performance status of at least 70. (4) Measurable disease on imaging. (5) At least 3 months since the completion of brain radiotherapy. (6) A lesion considered resectable at repeat resection.

Main exclusion criteria were as follows. (1) Prior or concurrent malignancy. (2) Contraindications to surgery or MRI. (3) Porphyria or soy allergy. (4) Severe renal or hepatic impairment. (5) Pregnancy.

Baseline characteristics, including dose, MGMT promoter status and follow-up, are summarized in Table 1.

### Procedures

#### Hiporfin administration

Hiporfin (hematoporphyrin derivative sodium) was given as a single intravenous infusion of 5 mg/kg before surgery, and the institutional protocol specifies a 40 to 72-hour interval between dosing and photodynamic therapy. A skin test preceded infusion when required, and no other photosensitizing agent was given.

#### Resection and fluorescence assessment

Standard craniotomy and microsurgical tumor resection were performed. After fluorescence-guided resection under blue-light excitation, the tumor cavity and its margins were inspected under blue-light excitation, and the operating surgeon recorded the presence or absence of fluorescence, the same two categories being used to label the corresponding tissue specimens. Descriptions of fluorescence intensity in the source records were pooled into the analysis as fluorescence present (Supplementary methods). All procedures used the same microscope system which is the ZEISS Kinevo 900 with BLUE 400 mode^23^. Blue-light excitation was 400–410 nm, and fluorescence detection was 620–710 nm.

#### Photodynamic therapy

Immediately after resection, the cavity was illuminated with a 630-nm laser at 150 J/cm² and 1 W^18^. When two cavities were treated in the same patient, each was illuminated separately at the same energy density. Illumination lasted 40 minutes to 1 hour. One to three named subregions were illuminated per cavity (Supplementary Table S1). All patients received light-avoidance instructions. These are described in the Supplementary methods.

#### Feasibility and safety

Feasibility was defined as completion of a single preoperative dose, FGR and immediate cavity PDT within one procedure, without abandonment or deferral. Operative, PDT and safety variables were extracted from the surgical and clinical records. The complete list is given in the Supplementary methods. Intraoperative video was classified frame by frame into white-light, blue-light and fluorescence scenes, as described in the Supplementary methods.

### Specimen sampling and reference standard

Tissue was sampled from fluorescent and non-fluorescent areas of tumor, cavity wall and peritumoral tissue, according to the surgeon’s assessment. Each specimen was placed in a separate container labeled with its anatomical site and fluorescence status, and frozen sections for intraoperative diagnosis were taken where indicated. Specimens were processed routinely into formalin-fixed paraffin-embedded blocks and stained with hematoxylin and eosin plus a per-block immunohistochemical panel. The reference standard was the written histopathological report issued by pathologists specializing in neuro-oncology. Each diagnosis was assigned to the same specimen that carried the fluorescence label. Antibodies, staining conditions, classification rules and the remaining pathological detail are given in the Supplementary methods. The pathologists were not blinded to the intraoperative fluorescence status, which is reported as a potential source of bias (STARD 2015 item 13b is not satisfied)^21^.

### Statistical analysis

#### Analysis set

Specimens were included in the accuracy analysis if they carried an explicit intraoperative fluorescence annotation. Frozen-section specimens were excluded, as were bulk specimens that were not containerized, dural specimens, and peritumoral or cavity-margin samples without a fluorescence label. The reason for exclusion of every specimen is listed in Supplementary Table S2. The two dural specimens that carried an explicit annotation were reintroduced in a prespecified sensitivity analysis. The operating surgeon knew the clinical history and the previous imaging of every patient when the fluorescence assessment was recorded, but not the histopathological outcome. Both correspond to how the index test is applied in practice (STARD 2015 item 13a)^21^. The numbers of specimens included, excluded and re-analyzed are reported in Results.

#### Index test and reference standard

Intraoperative fluorescence recorded by the operating surgeon served as the index test, and the corresponding histopathological diagnosis served as the reference standard. Each specimen was classified as fluorescent or non-fluorescent and as tumor or non-tumor. Several specimens were obtained per patient, so the patient was the unit of analysis. Specimen-level crude estimates with Wilson 95% confidence intervals are reported descriptively only.

#### Bivariate random-effects model

Diagnostic accuracy was pooled using the bivariate random-effects model of Reitsma et al, equivalent to the hierarchical summary ROC model of Rutter and Gatsonis^24,25^. The two-level model structure and its full specification are given in the Supplementary methods (Supplementary Figure S1). Between-patient heterogeneity was summarized by the I² statistic, and the AUC was obtained from a summary ROC curve.

#### Between-patient correlation

The between-patient variance components were estimated at essentially zero. The correlation is therefore not reliably estimable from six patients, and its point estimate is not interpreted. Pooling was repeated with univariate random-effects models, equivalent to assuming ρ = 0. Those estimates are reported in Results.

#### Software

Analyses used R 4.6.1 with mada 0.5.12 and metafor 5.0.1. The analysis script, the per-patient two-by-two tables and the model outputs are provided as supplementary material.

#### Exploratory morphology

Whether fluorescence-positive non-neoplastic specimens differed from tumor on morphology alone was examined in an exploratory analysis of whole-slide images with frozen pathology foundation-model encoders. No model was trained and the analysis has no clinical intended use. The methods are given in the Supplementary methods (Supplementary Table S3, Supplementary Table S4 and Supplementary Table S5).

## Results

### Patient characteristics and treatment

#### Patients and tumors

Six patients completed both fluorescence-guided resection and intraoperative PDT procedures including four women and two men. All six underwent FGR and PDT between September and October 2024 (Table 1). Median age was 53.5 years, and the age range was 20 to 61 years. All tumors were isocitrate dehydrogenase (IDH) wild-type glioblastoma, WHO CNS grade 4^22^. MGMT promoter methylation was determined molecularly in four patients. C02, C03 and C06 were methylated, C04 was unmethylated, and in C01 and C05 the status was inferred from MGMT immunohistochemistry (Table 1). Ki-67 indices reported in the diagnostic specimens ranged from 5% to 60% across the six patients (Table 1). C02 harbored two anatomically distinct lesions, a recurrent left parieto-occipital lesion in the previous surgical cavity and a newly diagnosed left temporal lesion. Both were resected during a single craniotomy and counted as two separate resection fields, so the cohort comprised six patients and seven resection fields. Surgeon-assessed gross total resection was achieved in all fields. Serial MRI of the two lesions resected in C02 is shown in Supplementary Figure S2. Paired preoperative and postoperative imaging of all six patients is shown in Supplementary Figure S3.

#### Dosing and intervals

Hiporfin was administered as a single intravenous infusion of 5 mg/kg before surgery. Body weight ranged from 46 to 80 kg and the administered dose from 230 to 400 mg (Table 1). The interval from the start of the infusion to skin incision was 40.0–46.3 hours, with a median of 45.6 hours. The interval to the start of illumination was 46.9–53.2 hours, with a median of 52.6 hours.

#### Specimen sampling

A total of 87 tissue specimens were submitted for histopathological examination, of which 63 from all six patients carried an explicit intraoperative fluorescence annotation and formed the primary analysis set (2 to 22 specimens per patient, Supplementary Table S6). The remaining 24 specimens were excluded — frozen sections, bulk specimens, dura and specimens without an explicit fluorescence annotation.

### Diagnostic accuracy

#### Primary analysis

Against histopathology, pooled sensitivity was 0.751 (95% CI 0.600–0.858) and specificity 0.606 (0.407–0.775) in the bivariate random-effects model, with an AUC of 0.739 (partial AUC 0.685). Crude positive and negative predictive values were 0.833 (0.694–0.917) and 0.619 (0.409–0.792). Of the 63 specimens, 35 were true positive, 7 false positive, 8 false negative and 13 true negative. The summary receiver operating characteristic (SROC) curve and the per-case estimates are shown in Figure 2A and 2B, and the cross-tabulation in Table 2. The crude specimen-level estimates were sensitivity 0.814 (0.674–0.903), specificity 0.650 (0.433–0.819) and accuracy 0.762 (0.644–0.850); between-patient heterogeneity was low (I² = 10.7%). Two patients (C01, C04) contributed no non-tumor specimen, so their specificity was undefined and they are not on the SROC curve.

**Figure 2.**
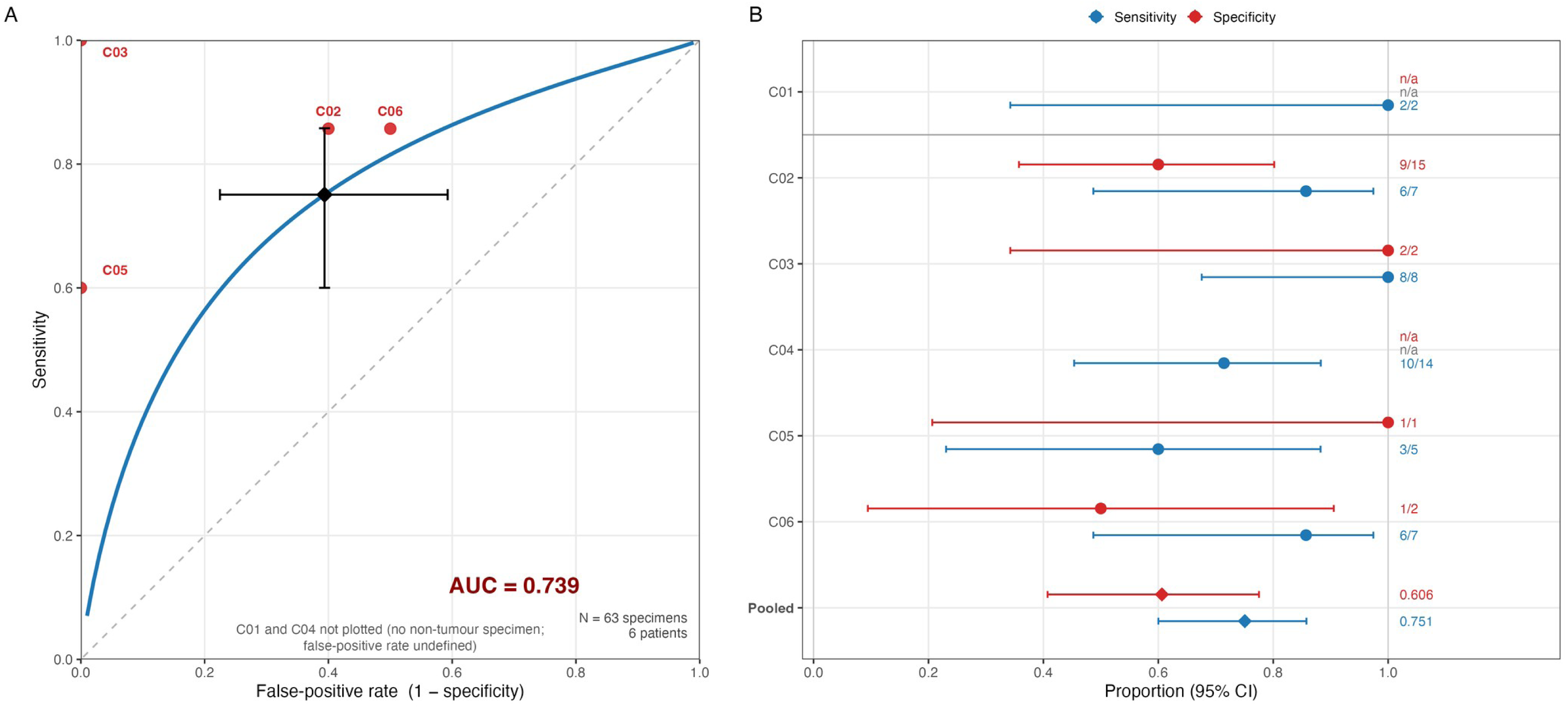
Diagnostic accuracy of intraoperative fluorescence against histopathology in 63 specimens from six patients. **A**, summary receiver operating characteristic curve derived from the bivariate random-effects model (solid line). The filled circles are the observed sensitivity and false-positive rate of the four patients from whom non-tumor specimens were obtained, labeled with their case identifiers; the open circle is the pooled estimate with its 95% confidence interval, and the dashed line is the line of no discrimination. The area under the curve is 0.739. No non-tumor specimen was obtained in the remaining two patients, so their false-positive rate is undefined and they do not appear in this panel. **B**, per-case sensitivity (blue) and specificity (red) with 95% confidence intervals; the numbers beside each estimate are the number of specimens classified correctly over the number examined, and n/a indicates that specificity is undefined because no non-tumor specimen was obtained. Pooled estimates (sensitivity 0.751, specificity 0.606) are shown separately at the bottom. ROC = receiver operating characteristic.

#### Sources of diagnostic error

All seven false-positive specimens were reported histopathologically as peritumoral tissue (not tumor), six of them from C02, and all eight false negatives were specimens recorded as non-fluorescent yet reported as glioma; the per-patient distribution is given in the Supplementary results. Representative histopathological images of the four fluorescence–pathology categories are shown in Figure 3.

**Figure 3.**
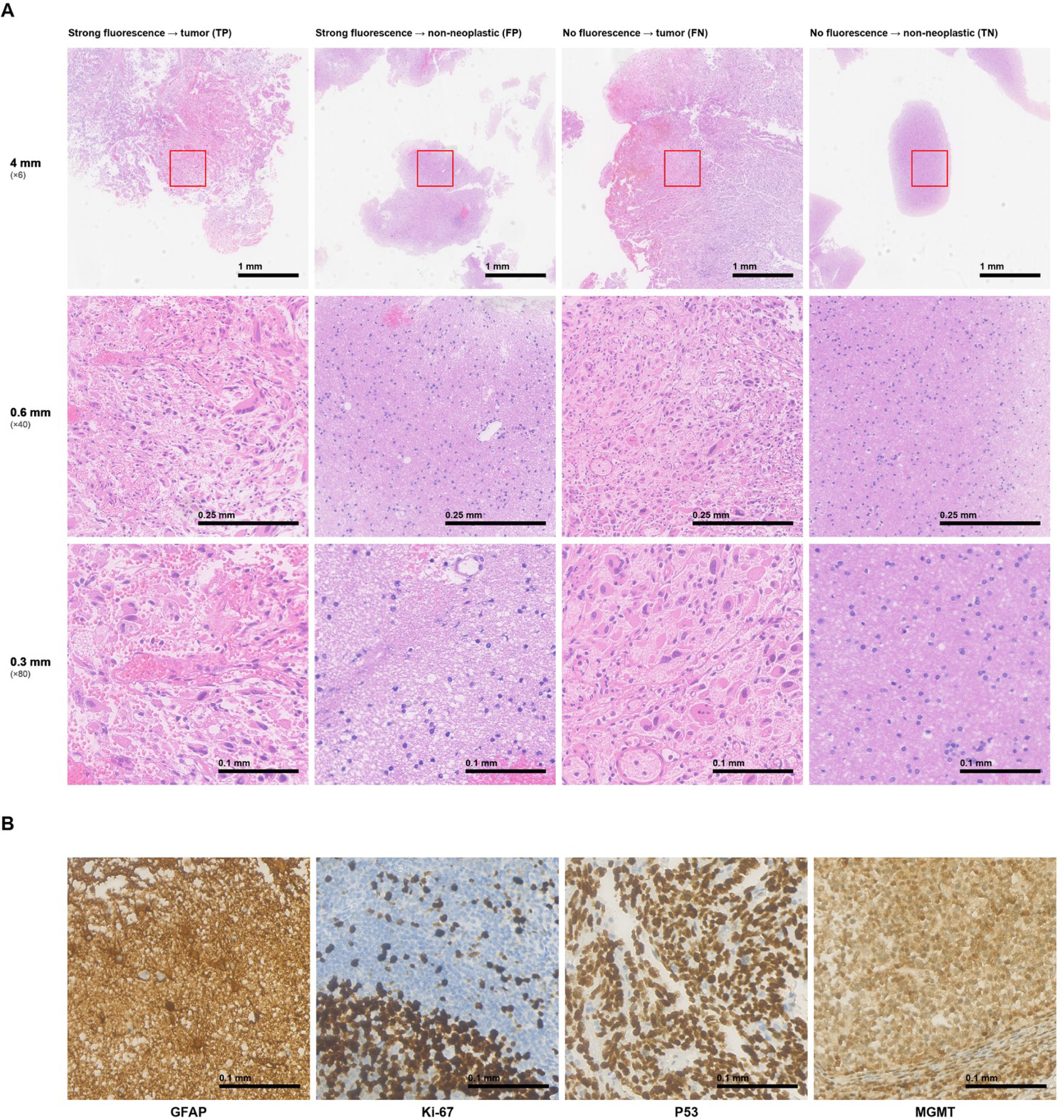
Histopathological findings according to intraoperative fluorescence status and final histopathological diagnosis (representative specimens from the patient who underwent resection of two lesions, C02). **A**, Representative hematoxylin and eosin sections from four specimens: strong fluorescence with tumor (true positive, specimen 2), strong fluorescence with non-neoplastic tissue (false positive, specimen 18), no fluorescence with tumor (false negative, specimen 10) and no fluorescence with non-neoplastic tissue (true negative, specimen 20). Each column shows the same specimen at three magnifications: a 4-mm field (×6), a 0.6-mm field (×40) and a 0.3-mm field (×80); red boxes mark the region shown at the next higher magnification. Scale bars, 1 mm (upper row), 0.25 mm (middle row) and 0.1 mm (lower row). All false-positive specimens in this series were reported histopathologically as peritumoral tissue. **B**, Immunohistochemical staining of tumor block 1A for glial fibrillary acidic protein (GFAP), Ki-67, p53 and O⁶-methylguanine-DNA methyltransferase (MGMT); scale bars, 0.1 mm. The Ki-67 indices reported with the diagnostic specimens are given in Table 1. MGMT staining was negative, consistent with the original pathology report. Block 1A is a diagnostic gross-specimen block and is not part of the 63-specimen analysis set; the immunohistochemical panels therefore illustrate the molecular phenotype of the tumor and are not tied to the fluorescence status of an individual specimen.

#### Sensitivity analyses

Excluding the two dura specimens that themselves carried a fluorescence annotation (analysis set n = 65) changed specificity from 0.606 to 0.574 (AUC 0.702) and left sensitivity unchanged (0.751). Leave-one-patient-out estimates (sensitivity 0.730–0.781, specificity 0.585–0.630, AUC 0.647–0.744) and the univariate random-effects refits are given in the Supplementary results; no single patient drove the pooled result.

#### Morphological comparison of false-positive specimens

On morphology alone, the false-positive slides were distinct from the tumor slides and indistinguishable from the non-neoplastic slides. Across the eight frozen pathology-encoder configurations, the cosine distance from the false-positive centroid to the true-positive centroid was 0.024 to 0.084 (p = 0.001 to 0.038). The distance to the false-negative centroid was 0.015 to 0.073 (p = 0.016 to 0.053). The distance to the true-negative centroid was much shorter, 0.002 to 0.024. No configuration separated the false-positive from the true-negative slides (p = 0.23 to 0.43). The false-positive specimens therefore behaved as non-neoplastic tissue and not as tumor (Supplementary Figure S4). The analysis used 76 HE-stained slides with each slide as the unit of analysis. The analysis set, every comparison including those that were not significant, and all robustness analyses are given in Supplementary Tables S3–S5.

#### Within-patient contrast (C02)

C02 had a newly diagnosed and a recurrent lesion resected in one procedure, so both fields were assessed with the same drug and illumination. In the newly diagnosed field, fluorescence identified both tumor specimens (2/2) and spared 7 of the 11 non-tumor specimens (7/11); all four false positives were peritumoral tissue and no tumor was missed. In the recurrent-cavity field, 4 of 5 tumor specimens and 2 of 4 non-tumor specimens were classified correctly, and the only false negative came from this field. Representative white-light and blue-light images of the same operative field are shown in Figure 4A and 4B, with additional exported frames in Supplementary Figure S5. The two fields were not statistically distinguishable, so this contrast is descriptive only.

**Figure 4.**
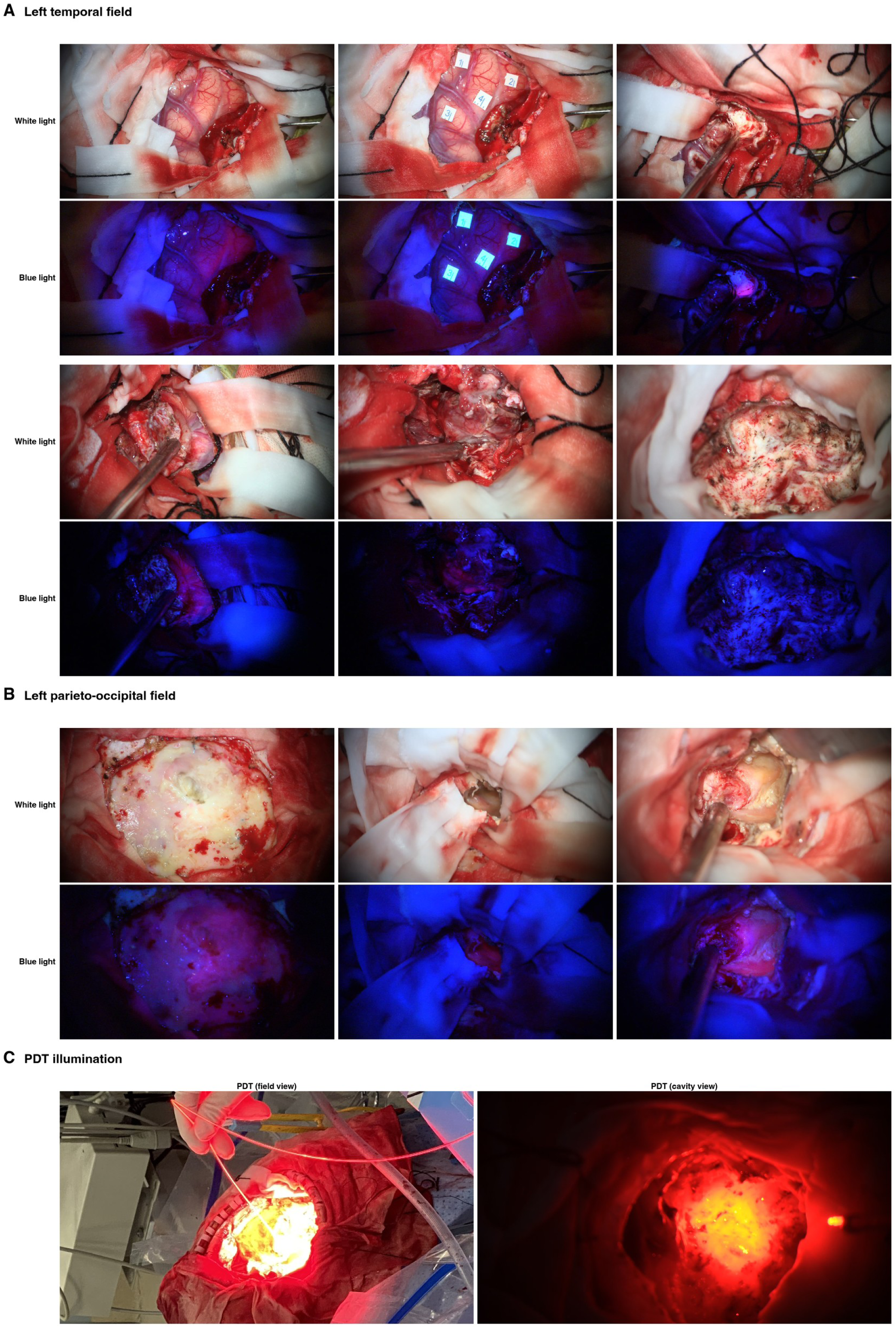
Intraoperative fluorescence imaging and photodynamic therapy in a patient with recurrent glioblastoma (C02). **A**, Left temporal field (six pairs of frames). **B**, Left parieto-occipital field (three pairs of frames). In **A** and **B**, each pair shows the same operative field under white light (upper image) and under 405-nm blue-light excitation for fluorescence (lower image); pairs are arranged from left to right in the order in which the fields were imaged. Fluorescent tissue appears magenta to red within the blue-light images. **C**, Illumination for photodynamic therapy (630 nm): field view (left) and cavity view (right). Frames that were exported during the analysis but not displayed here are provided in Supplementary Figure S5.

### Feasibility and safety

#### Completion and observation time

All six patients completed the full sequence of fluorescence-guided resection and photodynamic therapy in a single procedure. No treatment was abandoned or deferred (7/7 resection fields; Figure 4C). Photodynamic therapy was delivered at 630 nm in all six patients; the energy, power, illumination time and illuminated subregions for every resection field are listed in Supplementary Table S1. Continuous video was available for five of the six patients (118 fragments, 829 minutes in total). Frame-level scene classification, validated against 96 surgeon-annotated segments, gave 151.5 minutes under blue-light excitation, 100.0 minutes of which showed tumor fluorescence. White-light periods accounted for the remaining 677.5 minutes (Supplementary Table S7).

#### Safety

Estimated intraoperative blood loss, recorded for all six patients, averaged 300 mL (range 200–400 mL). Only C02 received a transfusion (200 mL of fresh frozen plasma). Somatosensory and motor evoked potentials remained stable. No new postoperative neurological deficit was documented. Postoperative complications were limited to one central nervous system infection (C02). The other five patients had an uneventful course. No clinically evident cutaneous photosensitivity was recorded. All patients received light-avoidance instructions (Supplementary Table S8).

## Discussion

In this prospective series, the hematoporphyrin derivative Hiporfin was used for fluorescence-guided resection of recurrent glioblastoma. To our knowledge, this is the first use of Hiporfin for intraoperative FGR in glioma quantified at the specimen level against histopathology. The study addressed diagnostic accuracy and procedural feasibility, not efficacy. Accuracy was moderate (AUC 0.739). The clinically important feature is the asymmetry. Fluorescent tissue was usually tumor, whereas non-fluorescent tissue could not be assumed to be tumor-free. Eight non-fluorescent specimens proved to be glioma, so fluorescence alone cannot declare a margin clear.

All seven false-positive specimens were peritumoral tissue, and six of them came from C02. In C02 the false-positive tissue came from both of the resected lesions, the newly diagnosed left temporal lesion and the recurrent lesion in the previously operated and irradiated cavity. Fluorescence-positive tissue that contains no tumor has been attributed to reactive astrocytosis with scattered inflammatory cells rather than to tumor^7,8^.

This pattern fits the reported difference between recurrent and newly diagnosed tumors. Our cohort was entirely recurrent, and 7 of the 20 tumor-free specimens were fluorescent. Pooled comparative data show the same direction at recurrence (10.1% versus 4.1%; OR 0.24, 95% CI 0.06–0.91)^11^. Specificity and negative predictive value are likewise lower for 5-ALA in recurrent tumors^26^.

An exploratory morphological analysis of the same specimens pointed in the same direction. With frozen pathology foundation-model encoders, the false-positive slides differed from the tumor slides in all eight configurations. They also differed from the false-negative slides in seven of eight configurations, but never from the non-neoplastic slides (Supplementary Figure S4). Fluorescent tissue that contained no tumor therefore resembled non-neoplastic tissue. It did not resemble the tumor that the surgeon overlooked. This is the pattern expected of peritumoral or reactive tissue, and it explains why accuracy here is limited by specificity.

In C02, accuracy was numerically higher in the newly diagnosed field on both measures. The difference was not statistically distinguishable with 13 and 9 specimens. The direction matches the reported fall in 5-ALA specificity at recurrence^26^. Hiporfin may therefore perform better in newly diagnosed glioblastoma. We are planning a prospective study of the same integrated workflow in that setting, with specimen-level accuracy against histopathology as the primary endpoint. The proportion of fluorescent non-tumor specimens will be compared with this cohort.

The only directly comparable strategy is 5-ALA fluorescence-guided resection followed by intraoperative open photodynamic therapy^12,13^. It was applied in 20 patients with recurrent high-grade glioma, and median progression-free survival was 6 months^12^. It established the dual-strategy concept, reported local control, and gave no diagnostic-accuracy estimate against histopathology. Our contribution is complementary. We addressed the same question with a different agent and provide specimen-level estimates of the diagnostic accuracy achieved by intraoperative visual assessment.

Published series that used histopathology as the reference standard are summarized in Supplementary Table S9^27-33^. Our values sit within that range. Two features of our series may explain this. In previously treated fields, the histopathological reference frequently read as peritumoral tissue or reactive change. Fluorescence was graded by eye without quantitative thresholds. Our morphological data point to visual grading as the limiting step.

HpD photodynamic therapy was reported for glioma surgery previously^18^. This phase I/II series treated 22 high-grade glioma patients after tumor resection, with HpD at 5 mg/kg and 630 nm illumination of the tumor bed. Hiporfin is a separate HpD preparation, but we also used the drug for photodynamic therapy. Our main innovation was to combine photodynamic therapy with fluorescence-guided resection. Hiporfin is a mixture of porphyrins, and its fluorescence is intrinsic to the drug^14^. After intravenous administration the porphyrins accumulate preferentially in tumor tissue relative to normal brain, and blue-light excitation shows them as red fluorescence^16,17^. 5-ALA works by a different route. 5-ALA is not fluorescent itself, and glioma cells convert it into protoporphyrin IX through the heme pathway^9,34^. That metabolic step is the source of its tumor selectivity. Hiporfin requires no such conversion, so its signal follows delivery and retention. The fluorescence used for guidance and the cytotoxic effect share a photochemical basis. Energy transfer from the excited triplet photosensitizer to molecular oxygen generates singlet oxygen, the Type II reaction common to most photosensitizers^16,17^. Our study added specimen-level evidence for Hiporfin-assisted fluorescence-guided resection of glioma. Therefore we offered a novel fluorescence-guided option in glioma surgery.

Our study has several limitations. Firstly, this is a small single-arm study of six patients and 63 specimens. It is hypothesis-generating and has no efficacy claim. Secondly, fluorescence by Hiporfin was a binary call determined by surgeon, so no intensity threshold could be examined. And the histopathological assessors were not blinded to the fluorescence status^21^. Finally, extent of resection was not verified volumetrically, and follow-up data was immature.

In recurrent glioblastoma, Hiporfin-guided fluorescence identified tumor with moderate accuracy. A fluorescent region is likely tumor. A non-fluorescent region still contains tumor. The error came from peritumoral tissue in the previously treated cavity. The complete workflow was feasible in every patient. Taken together, our study provides Hiporfin as a new option for fluorescence-guided glioma surgery and a potential tool for improving the extent of resection and local control.

## Supporting information

STRAD 2015 Checklist

## Acknowledgements

This resource was supported by the National Key R&D Program of China (No. 2023YFA1008600, to Z.H.), the Key Science and Technology Project of Guangdong Province, China (No. 2023B0303020002, to Y.M.), The National Natural Science Foundation of China (No. 82373386 to Y.M.), the National Natural Science Foundation of China (No. 82203129, to Z.H.) and the Guangdong Basic and Applied Basic Research Foundation (No. 2023A1515012509, to Z.H.). Hiporfin was supplied by Chongqing Huading Modern Biopharmaceutical Co., Ltd. as non-financial support. The company had no role in the design, conduct, analysis, or reporting of this study.

## Ethics approval and consent to participate

This study was approved by the Ethics Committee of Sun Yat-sen University Cancer Center, which serves as the Institutional Review Board of Sun Yat-sen University Cancer Center (651 Dongfeng Road East, Guangzhou 510060, China). Decision: ethical approval was granted (approval No. B2021-199-01; amendments Y01, Y02 and X01). Written informed consent, including consent for the research use of de-identified operative video and pathology material, was obtained from every patient. The study was conducted in accordance with the Declaration of Helsinki.

## Conflict of interest

The authors report no conflict of interest concerning the materials or methods used in this study or the findings specified in this paper.

## Author contributions

Conception and design: Y.M., X.Z. Acquisition of data: C.W., Z.H., W.H., L.W., H.D., Y.C. Analysis and interpretation of data: Z.H., G.Z. Drafting the article: Z.H. Critically revising the article: all authors. Reviewed and approved the final version: all authors. Agreed to be accountable for all aspects of the work: all authors.

## Data availability

De-identified case-level data are available from the corresponding author on reasonable request. Raw operative video and whole-slide pathology images are not publicly available because of patient privacy and ethical restrictions but can be reviewed under ethics approval and institutional permission. The study protocol is registered at ClinicalTrials.gov (NCT07832448; STARD 2015 item 29).

## Use of artificial intelligence

Generative artificial intelligence tools were used to improve the language and readability of the manuscript. They were not used to generate, alter or select research data, and no artificial intelligence tool is an author or is credited with authorship. The exploratory morphological comparison used publicly available pathology foundation-model encoders as frozen feature extractors; no model was trained, and no output informed any clinical decision. The authors take full responsibility for the content of the manuscript.

---

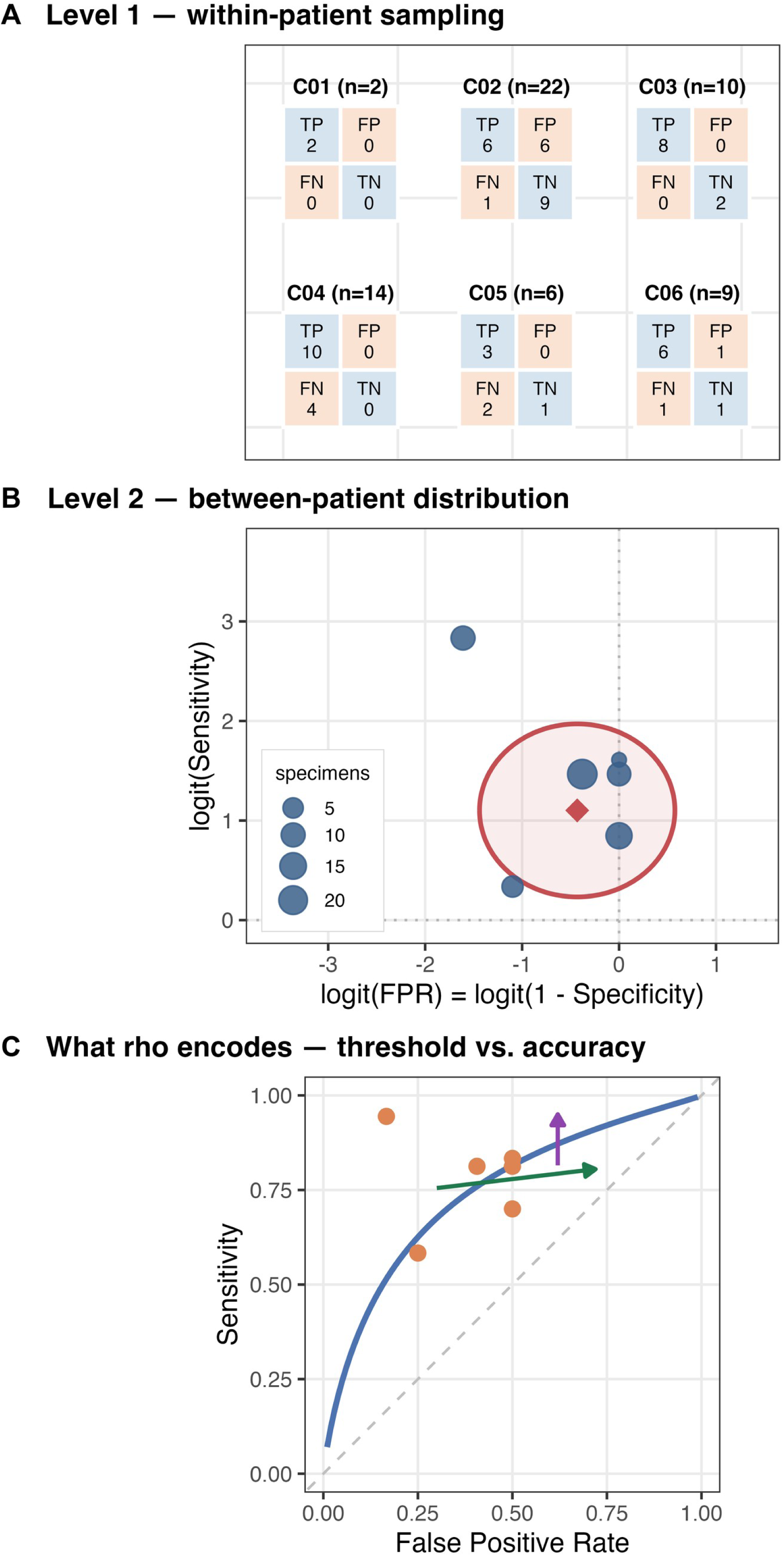

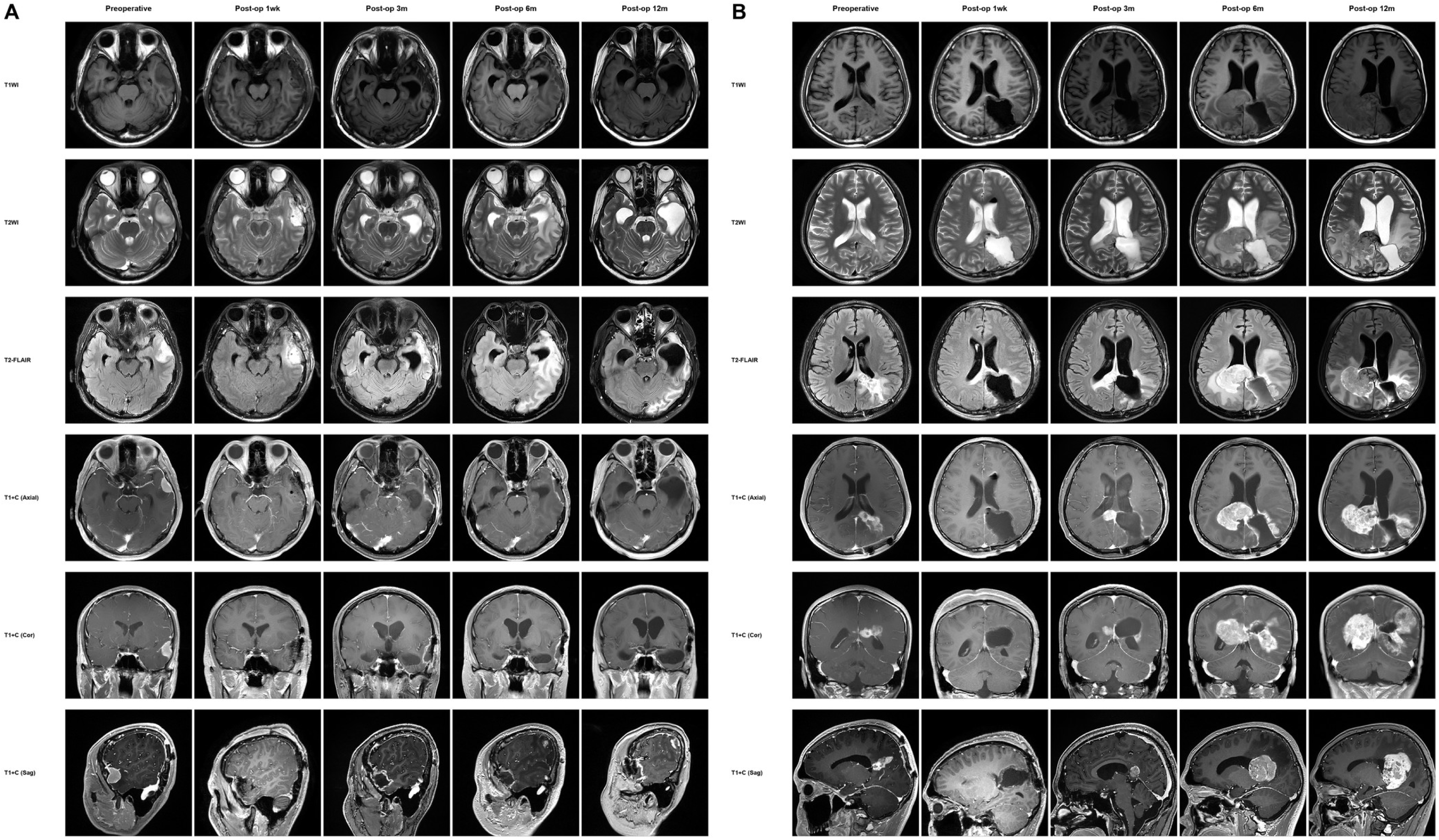

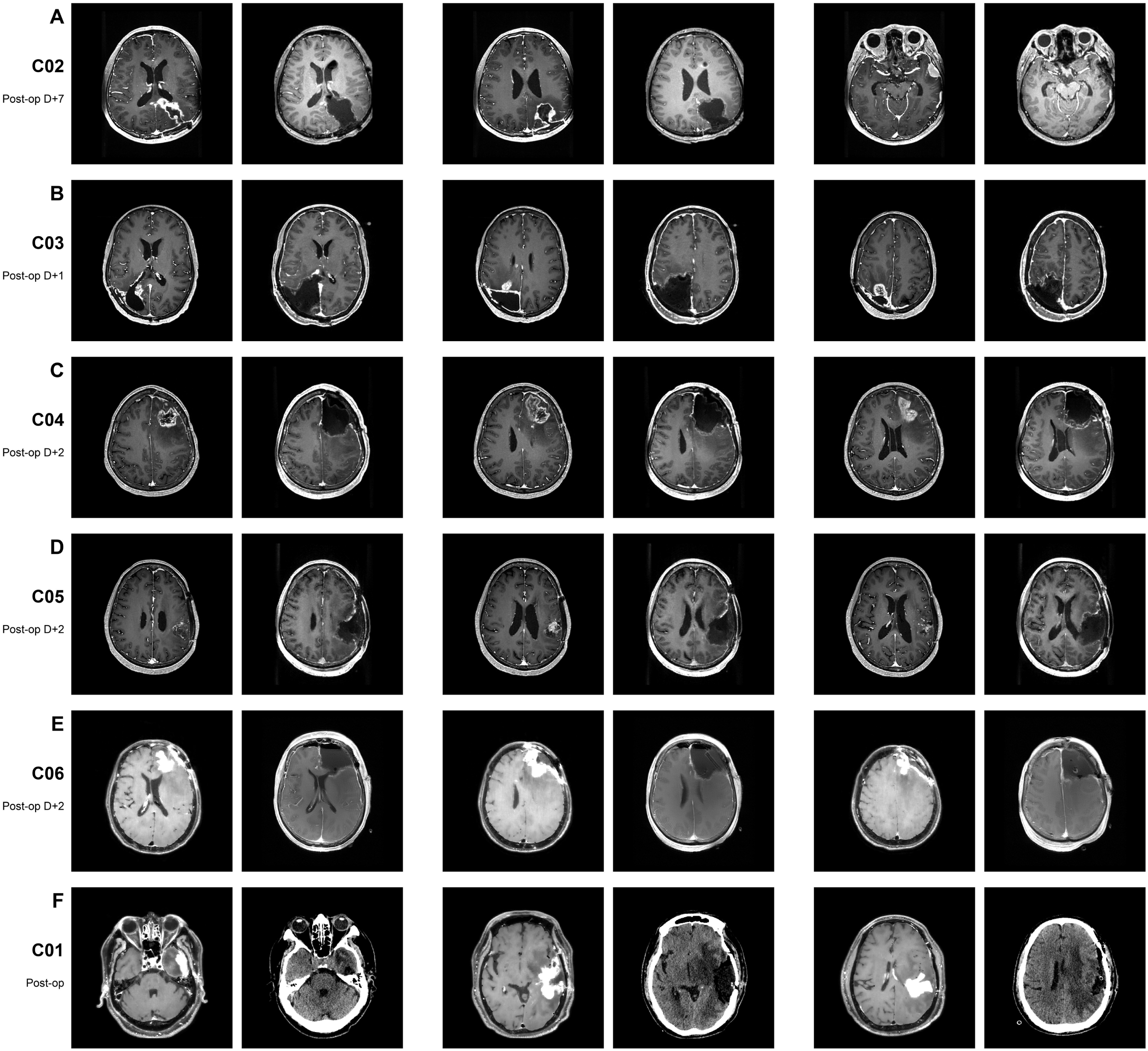

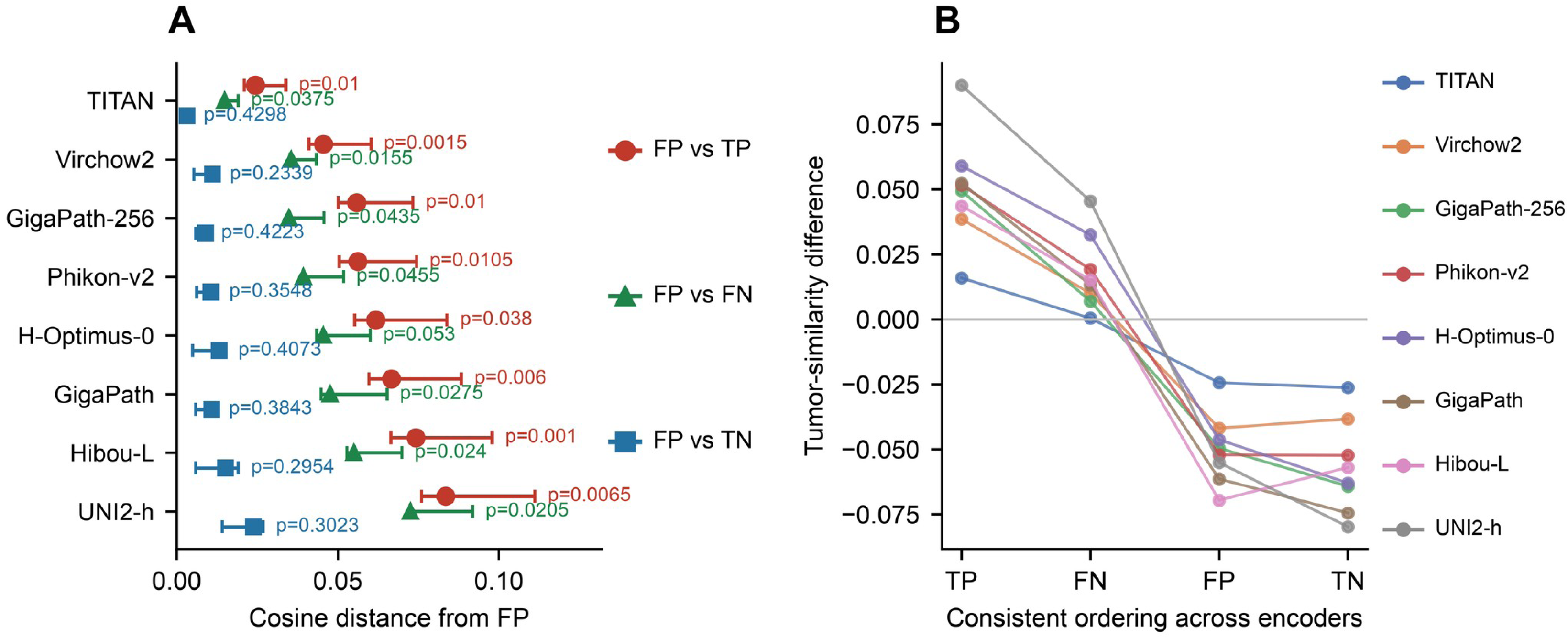

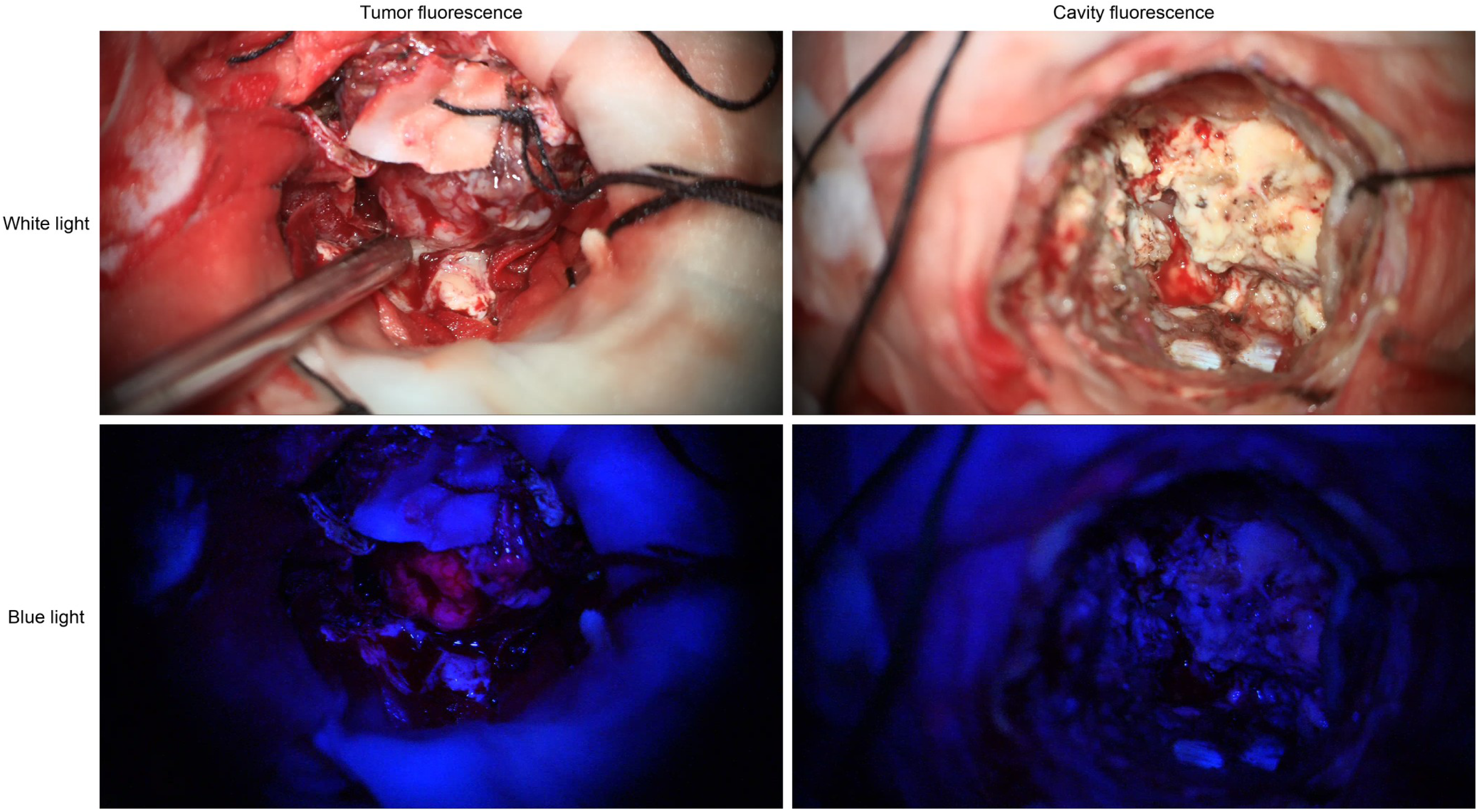

## Notes

### Competing Interest Statement

The authors have declared no competing interest.

### Clinical Trial

NCT07832448

### Author Declarations

The Ethics Committee of Sun Yat-sen University Cancer Center (Institutional Review Board of Sun Yat-sen University Cancer Center), 651 Dongfeng Road East, Guangzhou 510060, Guangdong Province, China, gave ethical approval for this work (approval number B2021-199-01; amendments Y01, Y02 and X01). Written informed consent, including consent for the research use of de-identified operative video and pathology material, was obtained from every patient. The study protocol is registered at ClinicalTrials.gov (NCT07832448). The study was conducted in accordance with the Declaration of Helsinki.

