## Supplementary_material for "Hiporfin (HpD) for fluorescence-guided resection of recurrent glioblastoma: diagnostic accuracy and feasibility of an integrated photodynamic strategy"

### Supplementary methods

#### Supplementary methods. Definitions and classification rules.

**Pooling of fluorescence grades.** Specimens graded as strongly, moderately or weakly fluorescent were pooled as test-positive. Specimens without fluorescence were pooled as test-negative. Intensity was judged by eye and the cohort was small.

**Reference-standard classification rules.** Specimens reported as high-grade glioma, as glioma, or as containing scattered tumor cells were classified as tumor. Specimens reported as peritumoral tissue, reactive hyperplasia, or without definite tumor were classified as non-tumor.

**Per-specimen adjudication and re-review.** In one patient (C03) the pathology report grouped specimens 2–13 in a single statement. Two specimens of that patient (6 and 9), both non-fluorescent, were adjudicated as non-tumor after per-specimen slide review and counted as true negatives. All seven fluorescence-positive but non-tumor specimens were re-reviewed by the department of pathology. The non-neoplastic classification was confirmed.

**Dura as a non-target.** Dura is not a target of fluorescence-guided resection. The two dural specimens that carried an explicit fluorescence annotation were reintroduced in a prespecified sensitivity analysis (n = 65).

**Protocol window check.** The interval to incision and the interval to illumination both lay within the protocol's 40–72-hour window for blue-light illumination in all six patients. Both intervals lay within its stricter 48–72-hour window in five patients. The interval to illumination was 46.9 hours in C06.

#### Supplementary methods. Immunohistochemical panel and microscope.

**Immunohistochemical panel.** The per-block panel comprised GFAP, Olig-2, IDH1, ATRX, p53, MGMT and Ki-67. Clone numbers and dilutions are not recorded in the source reports.

**Microscope.** All six procedures used a ZEISS Kinevo 900 in BLUE 400 fluorescence mode. The excitation was 400–410 nm and the detection range 620–710 nm. The imaging platform was identified from the intraoperative video recordings, because the operative notes did not record it.

#### Supplementary methods. Operative, photodynamic-therapy and safety variables.

**Variables extracted.** Estimated intraoperative blood loss, transfusion, intraoperative somatosensory and motor evoked potentials where monitored, postoperative complications, new postoperative neurological deficits, and light-avoidance instructions with any documented cutaneous photosensitivity.

**Photodynamic therapy parameters recorded per resection field.** Wavelength, energy density or absolute energy, power, illumination duration and the named illuminated subregions (Supplementary Table S1).

**Light-avoidance instructions.** Ranging from environmental light avoidance to avoidance of strong light sources, including direct sunlight, for one month.

#### Supplementary methods. Video recording and scene classification.

Intraoperative video was recorded continuously whenever the recording system was in use. Five of six patients were recorded, in 118 fragments and 829 minutes in total. Each frame was classified from its color-channel statistics after decoding the recordings over their full range. A blue-light scene was defined by a blue-channel fraction of at least 5%. A frame within a blue-light scene was classified as showing tumor fluorescence when the magenta fraction was at least 2.0% and no focusing-light core was present. The thresholds were validated in one patient against 96 segments annotated by the operating surgeon. Frame-level sensitivity was 0.833 and specificity 0.865 (2,952 frames). Segment-level sensitivity was 0.885 and specificity 0.933, with a segment-clustered bootstrap interval of 0.7 to 3.2 percentage points around the threshold. The resulting observation times are given in Supplementary Table S7. The earlier timing values obtained with an incorrect color-range decoding setting were discarded. The full recomputation is described in the project record.

### Supplementary methods. Bivariate model specification.

Patient-level true-positive and false-positive counts were modeled with binomial likelihoods. Patients therefore contribute in proportion to the number of specimens examined. The patient-specific logit pairs (logit Se, logit FPR) were assumed to follow a bivariate normal distribution with mean ($\mu$_Se, $\mu$_FPR) and an unstructured $2 \times 2$ between-patient covariance matrix $\Sigma$. Variance components were estimated by restricted maximum likelihood. A continuity correction of 0.5 was applied to all cells, because two patients contributed no non-tumor specimen. The correlation between logit sensitivity and logit false-positive rate distinguishes patients differing along the threshold axis from those differing in overall accuracy. Along the threshold axis both measures rise together ($\rho > 0$). A rise in sensitivity at a fixed false-positive rate indicates overall accuracy ($\rho < 0$).

**Within-patient correlation.** Within-patient correlation is not modeled. The between-patient variance components were estimated at essentially zero ($4.0 \times 10^{-5}$ on the logit sensitivity scale and $4.5 \times 10^{-5}$ on the logit false-positive-rate scale). The bivariate model therefore reduced to fixed effects.

### Supplementary methods. Exploratory morphological analysis of histopathological specimens.

**Analysis set.** We examined whether specimens that were fluorescent but non-neoplastic differed from tumor on morphology alone. The available whole-slide images were analyzed in an exploratory manner. Of 246 slides, 76 HE-stained slides carried both an intraoperative fluorescence status and a histopathological diagnosis and were included. The analysis set comprised 42 true-positive, 15 true-negative, 12 false-negative and 7 false-positive slides from six patients and 64 blocks (Supplementary Table S3). The slide was the unit of analysis. No model was trained, no diagnostic performance is reported, and no result was used for a clinical decision.

**Encoders and descriptor.** Tissue was detected on the pyramid summary image. Tiles of $512 \times 512$ pixels at 0.5 µm/px (20×) were sampled on a jittered grid within the tissue mask, with an identical tile coordinate set for every model. Eight frozen configurations were applied to those tiles. Seven were distinct tile-level encoders: Virchow2, Phikon-v2, H-Optimus-0, Hibou-L, UNI2-h, the GigaPath tile encoder and the TITAN/CONCH v1.5 patch encoder. GigaPath was also run at its native 256-pixel field size. The TITAN slide encoder, which learns its own spatial aggregation, was used as a robustness check. Weights were loaded with strict key matching and verified file by file against the published hashes. Dimensions, revisions and license terms are listed in Supplementary Table S4. Four configurations are permissively licensed and four (Phikon-v2, UNI2-h, Virchow2 and TITAN) are restricted to non-commercial research. All were used exclusively for research. The descriptor of a slide was the L2-normalized mean of its tile feature vectors. For the slide encoder the model's own slide vector was used.

**Statistical analysis.** Groups were compared by the cosine distance between group centroids (2,000 label permutations). Every comparison was repeated with leave-one-case-out resampling, in which one patient is removed at a time. Because slides are nested within patients, the nominal p values are not used as the summary statistic. The intervals are the conservative summary, and they are reported with every comparison. A one-dimensional axis, cos(slide, true-positive centroid) − cos(slide, true-negative centroid), expressed morphological similarity to tumor relative to non-neoplastic tissue. Patients contributing slides to both groups were also compared internally. Preparatory quality was examined by correlating the first two principal components of the slide descriptors with tissue fraction, slide area, gray value and tissue RGB values. Predicted multiplex immunofluorescence (23 channels) generated from the same slides served as a negative control experiment (Supplementary Table S4 and Supplementary Table S5). Thirty-two nominal tests were performed without correction for multiple comparisons. The strength of the evidence rests on directional concordance across independently pretrained encoders. No single p value is decisive.

### Supplementary results

#### Supplementary results. Per-patient distribution of diagnostic errors.

The pooled analysis set comprised 35 true-positive, 7 false-positive, 8 false-negative and 13 true-negative specimens (Table 2). The per-case classification of the 63 analysis-set specimens is also given in Table 2. No non-tumor specimen was obtained in two patients (C01 and C04), and their specificity is therefore undefined. The seven false-positive specimens came from two patients: six in C02 and one in C06. The eight false-negative specimens came from four patients: four in C04, two in C05, and one each in C02 and C06. The reported histopathological wording and the reference call for every specimen are given in Supplementary Table S6.

#### Supplementary results. Sensitivity analyses.

**Leave-one-patient-out and univariate refits.** Leave-one-patient-out analysis gave sensitivity 0.730–0.781, specificity 0.585–0.630 and AUC 0.647–0.744. All intervals overlapped the primary estimate. Univariate random-effects refits led to the same conclusion. No single patient drove the pooled result.

#### Supplementary results. Morphological comparison of false-positive specimens.

**Morphological comparison of false-positive specimens (exploratory).** We examined whether false-positive specimens differed from tumor on morphology alone. Tile-level features (512 × 512 px at 0.5 µm/px, identical coordinates across encoders) were extracted from the available whole-slide images. Group centroids were compared using eight frozen pathology-encoder configurations (seven distinct tile-level encoders, one of which was also evaluated at its native field size). The cosine distance between false-positive and true-positive specimens was significant for every encoder ($p = 0.001$–$0.038$ by permutation testing). Its leave-one-case-out ranges were 0.021–0.111, and none included zero. The distance between false-positive and false-negative specimens was significant in seven of eight encoders (H-Optimus-0, $p = 0.053$). The distance between false-positive and true-negative specimens was not significant for any encoder ($p = 0.234$–$0.430$). The tumor-similarity difference was defined as cosine similarity to the true-positive centroid minus cosine similarity to the true-negative centroid. It was positive in true-positive specimens, intermediate in false-negative specimens and negative in false-positive and true-negative specimens. This ordering was consistent across all eight configurations (Supplementary Figure S4). The difference between false-positive and true-negative specimens was in the same direction in all 18 within-patient comparisons available. The false-positive specimens were therefore closest to non-neoplastic tissue, yet distinguishable from the false-negative specimens that contain tumor.

The analysis included 42 true-positive, 15 true-negative, 12 false-negative and 7 false-positive HE-stained slides, 76 slides in total, and used each slide as the unit of analysis. Slide numbers therefore differ from the 63-specimen analysis set. The analysis was exploratory and tile-level, and the encoders were used without any task-specific training. It is presented as supporting morphological evidence and not as a diagnostic model. Nominal p values are uncorrected for multiple comparisons. Supplementary Table S4 gives every comparison, dimension, revision and license term, and Supplementary Table S3 gives the analysis set.

### Supplementary tables

#### Supplementary Table S1. Photodynamic therapy illumination parameters

| Case | Wavelength | Energy density | Power | Illumination time | Interval to PDT, h | Illumination regions |
|---|---|---|---|---|---|---|
| C01 | 630 nm | 150 J/cm² | 0.8–1.5 W | 1 h | 53.2 | 3 |
| C02 | 630 nm | 150 J/cm² | 1 W | 40 min | 52.7 | **2** |
| C03 | 630 nm | 150 J/cm² | 1 W | 40 min | 52.5 | 1 |
| C04 | 630 nm | 150 J/cm² | 1 W | 40 min | 53.2 | 1 |
| C05 | 630 nm | 150 J/cm² | 1 W | 40 min | 52.4 | 2 |
| C06 | 630 nm | 150 J/cm² | 1 W | 1 h | 46.9 | 3 |

Footnotes

PDT, photodynamic therapy.

**Supplementary Table S2. Specimens excluded from the analysis set (n = 24)**

| # | Case | Specimen | Specimen name (as reported) | Fluorescence | Reason for exclusion |
|---|---|---|---|---|---|
| 1 | C01 | 2 | left temporal tumor (frozen 1) | Frozen section | Frozen-section specimen |
| 2 | C01 | 3 | left temporal tumor (frozen 2) | Frozen section | Frozen-section specimen |
| 3 | C01 | 4 | left temporal tumor (frozen 3) | Frozen section | Frozen-section specimen |
| 4 | C01 | 1 | left temporoparietal tumor, bulk | Bulk | Bulk specimen (not containerized) |
| 5 | C01 | 7 | peritumoral tissue | Peritumoral | Peritumoral tissue (no fluorescence label) |
| 6 | C02 | Frozen block 2 | left parieto-occipital corpus callosum (fluorescence) | Frozen section | Frozen-section specimen |
| 7 | C02 | 1 | left temporal tumor, bulk | Bulk | Bulk specimen (not containerized) |
| 8 | C02 | 30 | periventricular tumor 3 | No label | Not fluorescence-labeled |
| 9 | C02 | 25 | left parieto-occipital dura | Dura | Dura |
| 10 | C02 | 12-16 | left temporal cavity margins (all walls) | No label | Cavity margin (no fluorescence label) |
| 11 | C02 | 17 | left parieto-occipital epidural tissue | Dura | Dura |
| 12 | C03 | 1 | right parietal dural tissue (weak fluorescence) | Weak fluorescence | Dura |
| 13 | C03 | 12 | right parietal tumor 1, bulk | No label | Not fluorescence-labeled |
| 14 | C03 | 13 | right parietal tumor 2, bulk | No label | Not fluorescence-labeled |
| 15 | C03 | Frozen block | right parietal tumor (post-frozen paraffin) | Frozen section | Frozen-section specimen |
| 16 | C04 | 1 | left frontal dura (fluorescence) | Fluorescent | Dura |
| 17 | C04 | 16 | left frontal tumor, bulk | Bulk | Bulk specimen (not containerized) |
| 18 | C05 | 7 | left frontal dura | Dura | Dura |
| 19 | C05 | 8 | left frontal tumor, bulk | Bulk | Bulk specimen (not containerized) |
| 20 | C05 | Frozen block | left frontal tumor surface, fluorescence 1 (post-frozen paraffin) | Frozen section | Frozen-section specimen |
| 21 | C06 | 10 | left frontal tumor, bulk | Bulk | Bulk specimen (not containerized) |
| 22 | C06 | 11 | peritumoral tissue | Peritumoral | Peritumoral tissue (no fluorescence label) |
| 23 | C06 | Frozen block 1 | left frontal tumor, intraoperative fluorescence 1 (post-frozen paraffin) | Frozen section | Frozen-section specimen |

| | | | | | |
|---|---|---|---|---|---|
| 24 | C06 | Frozen block 2 | left frontal tumor, intraoperative fluorescence 2 (post-frozen paraffin) | Frozen section | Frozen-section specimen |

### Supplementary Table S3. Analysis set of the exploratory morphological comparison

#### A. Construction of the analysis set

| Step | Slides |
|---|---|
| Slides in the whole-slide image archive | 246 |
| Without an intraoperative fluorescence status and histopathological diagnosis (excluded from the labeled set) | 133 |
| With no histopathological annotation | 17 |
| With a paired fluorescence status and histopathological diagnosis | 96 |
| stained other than with HE (immunohistochemistry: Ki-67 12, GFAP 1, Olig-2 1, IDH1 1, ATRX 1, p53 1, MGMT 2, Ag 1) | 20 |
| HE-stained slides analyzed | 76 |
| Blocks represented | 64 |

#### B. Slides per patient and group

| Patient | true-positive | true-negative | false-positive | false-negative | total |
|---|---|---|---|---|---|
| C01 | 2 | 0 | 0 | 0 | 2 |
| C02 | 7 | 9 | 6 | 1 | 23 |
| C03 | 13 | 4 | 0 | 0 | 17 |
| C04 | 11 | 0 | 0 | 8 | 19 |
| C05 | 3 | 1 | 0 | 2 | 6 |
| C06 | 6 | 1 | 1 | 1 | 9 |
| **Total slides** | **42** | **15** | **7** | **12** | **76** |
| **Effective number of patients** | **6** | **4** | **2** | **4** | — |

Footnotes

HE, hematoxylin and eosin; IHC, immunohistochemistry; GFAP, glial fibrillary acidic protein; Olig-2, oligodendrocyte transcription factor 2; IDH1, isocitrate dehydrogenase 1; ATRX, alpha-thalassemia/mental retardation syndrome X-linked; MGMT, O6-methylguanine-DNA methyltransferase; Ki-67, marker of proliferation Ki-67.

### Supplementary Table S4. Exploratory morphological comparison — full encoder-level results

#### A. Group comparisons (cosine distance between centroids)

| Encoder | Model / descriptor | Dimension | License | FP vs TP distance | p | LOO range | FP vs FN distance | p | LOO range | FP vs TN distance | p | LOO range | FN vs TP distance | p | LOO range | FN vs TN distance | p | LOO range |
|---|---|---|---|---|---|---|---|---|---|---|---|---|---|---|---|---|---|---|
| TITAN (patch) | MahmoodLab/TITAN, CONCH v1.5 patch encoder | 768 | CC-BY-NC-ND-4.0 | 0.0244 | 0.01 | 0.021–0.0339 | 0.0149 | 0.0375 | 0.0176–0.0191 | 0.0033 | 0.4298 | 0.0023–0.0044 | 0.0084 | 0.069 | 0.007–0.012 | 0.0109 | 0.0455 | 0.0085–0.0217 |
| Virchow2 | paige-ai/Virchow2 | 2560 | CC-BY-NC-ND-4.0 | 0.0455 | 0.0015 | 0.041–0.0603 | 0.0355 | 0.0155 | 0.0352–0.0434 | 0.0111 | 0.2339 | 0.0054–0.0125 | 0.0104 | 0.1789 | 0.0078–0.0135 | 0.0173 | 0.0825 | 0.0161–0.0367 |
| GigaPath-256 | prov-gigapath/prov-gigapath (native 256-px field) | 384 | Apache-2.0 | 0.0558 | 0.01 | 0.0501–0.0733 | 0.0348 | 0.0435 | 0.0348–0.0458 | 0.009 | 0.4223 | 0.0059–0.0098 | 0.0203 | 0.089 | 0.0197–0.027 | 0.0253 | 0.0505 | 0.0241–0.0492 |
| Phikon-v2 | owkin/phikon-v2 | 1024 | Owkin non-commercial | 0.0562 | 0.0105 | 0.0505–0.0745 | 0.0394 | 0.0455 | 0.0395–0.0518 | 0.0106 | 0.3548 | 0.0062–0.0123 | 0.0158 | 0.1774 | 0.0139–0.0221 | 0.0259 | 0.078 | 0.0279–0.052 |
| H-Optimus-0 | bioptimus/H-optimus-0 | 1536 | Apache-2.0 | 0.0618 | 0.038 | 0.0552–0.084 | 0.0455 | 0.053 | 0.0435–0.0601 | 0.0133 | 0.4073 | 0.005–0.0151 | 0.0242 | 0.1589 | 0.0239–0.0315 | 0.035 | 0.0565 | 0.0369–0.0616 |
| GigaPath | prov-gigapath/prov-gigapath | 384 | Apache-2.0 | 0.0666 | 0.006 | 0.0598–0.0884 | 0.0476 | 0.0275 | 0.0448–0.0653 | 0.0109 | 0.3843 | 0.0059–0.0122 | 0.0234 | 0.091 | 0.0226–0.0301 | 0.0311 | 0.049 | 0.027–0.0614 |
| Hibou-L | histai/hibou-L | 1024 | Apache-2.0 | 0.0742 | 0.001 | 0.0665–0.098 | 0.055 | 0.024 | 0.0529–0.0699 | 0.0152 | 0.2954 | 0.0059–0.0191 | 0.0144 | 0.1459 | 0.0124–0.0206 | 0.0254 | 0.1134 | 0.0273–0.0663 |
| UNI2-h | MahmoodLab/UNI2-h | 1536 | CC-BY-NC-ND-4.0 | 0.0835 | 0.0065 | 0.076–0.1112 | 0.0726 | 0.0205 | 0.0725–0.0919 | 0.0238 | 0.3023 | 0.0142–0.0268 | 0.0286 | 0.1084 | 0.0286–0.0354 | 0.0548 | 0.0285 | 0.054–0.0938 |
| **TITAN (slide)** | MahmoodLab/TITAN, slide encoder | 768 | CC-BY-NC-ND-4.0 | 0.1056 | 0.003 | 0.0924–0.1391 | 0.034 | 0.1139 | 0.0359–0.0376 | 0.0867 | 0.001 | 0.078–0.1058 | 0.1105 | 0.0005 | 0.0782–0.2383 | | | |

**B. Tumor-similarity difference cos(slide, TP centroid) − cos(slide, TN centroid)**

| Encoder | TP median | TP IQR | TN median | TN IQR | FP median | FP IQR | FN median | FN IQR | FP vs TN p |
|---|---|---|---|---|---|---|---|---|---|
| TITAN (patch) | 0.0159 | -0.0008~0.0454 | -0.0263 | -0.0326~-0.0095 | -0.0244 | -0.0296~-0.0105 | 0.0004 | -0.0202~0.0192 | 0.971 |
| Virchow2 | 0.0385 | -0.0057~0.0691 | -0.0383 | -0.0542~-0.0173 | -0.0419 | -0.0453~-0.0232 | 0.01 | -0.0238~0.0200 | 0.8541 |
| GigaPath-256 | 0.0494 | 0.0052~0.0944 | -0.0642 | -0.0711~-0.0225 | -0.0496 | -0.0611~-0.0314 | 0.007 | -0.0405~0.0367 | 0.96 |
| Phikon-v2 | 0.0517 | -0.0150~0.0945 | -0.0523 | -0.0740~0.0089 | -0.0521 | -0.0681~-0.0138 | 0.0192 | -0.0529~0.0533 | 0.7941 |
| H-Optimus-0 | 0.059 | -0.0185~0.1113 | -0.063 | -0.0799~-0.0112 | -0.0463 | -0.0673~-0.0269 | 0.0325 | -0.0439~0.0425 | 0.9045 |
| GigaPath | 0.0524 | -0.0014~0.1075 | -0.0745 | -0.0829~-0.0148 | -0.0614 | -0.0714~-0.0417 | 0.0133 | -0.0508~0.0458 | 0.8771 |
| Hibou-L | 0.0436 | -0.0186~0.0913 | -0.057 | -0.0802~0.0183 | -0.0696 | -0.0798~-0.0285 | 0.0151 | -0.0584~0.0507 | 0.4183 |
| UNI2-h | 0.09 | 0.0005~0.1289 | -0.0798 | -0.1112~-0.0372 | -0.0551 | -0.0912~-0.0292 | 0.0455 | -0.0377~0.0568 | 0.6777 |
| TITAN (slide) | 0.1568 | 0.0900~0.1949 | -0.1655 | -0.1946~-0.1143 | -0.1329 | -0.1409~0.0023 | -0.0191 | -0.0577~0.0775 | 0.0195 |

**C. Within-patient comparisons (only patients contributing slides to both groups)**

| Encoder | Patient | FP slides | TN slides | FP median similarity difference | TN median similarity difference | difference (FP − TN) |
|---|---|---|---|---|---|---|
| TITAN (patch) | C02 | 6 | 9 | -0.0253 | -0.0296 | 0.0042 |
| TITAN (patch) | C06 | 1 | 1 | -0.008 | -0.0263 | 0.0183 |
| Virchow2 | C02 | 6 | 9 | -0.0428 | -0.0535 | 0.0108 |
| Virchow2 | C06 | 1 | 1 | -0.0059 | -0.0292 | 0.0234 |
| GigaPath-256 | C02 | 6 | 9 | -0.0537 | -0.071 | 0.0174 |
| GigaPath-256 | C06 | 1 | 1 | -0.0117 | -0.0384 | 0.0267 |
| Phikon-v2 | C02 | 6 | 9 | -0.057 | -0.0672 | 0.0102 |
| Phikon-v2 | C06 | 1 | 1 | -0.0172 | -0.0456 | 0.0284 |
| H-Optimus-0 | C02 | 6 | 9 | -0.0559 | -0.0785 | 0.0226 |
| H-Optimus-0 | C06 | 1 | 1 | -0.003 | -0.0447 | 0.0417 |
| GigaPath | C02 | 6 | 9 | -0.0634 | -0.082 | 0.0186 |
| GigaPath | C06 | 1 | 1 | -0.0083 | -0.0446 | 0.0363 |
| Hibou-L | C02 | 6 | 9 | -0.0733 | -0.0798 | 0.0065 |
| Hibou-L | C06 | 1 | 1 | -0.0234 | -0.0524 | 0.0289 |
| UNI2-h | C02 | 6 | 9 | -0.0709 | -0.1078 | 0.0369 |
| UNI2-h | C06 | 1 | 1 | 0.0082 | -0.0423 | 0.0504 |
| TITAN (slide) | C02 | 6 | 9 | -0.1352 | -0.1927 | 0.0575 |
| TITAN (slide) | C06 | 1 | 1 | -0.0129 | -0.1202 | 0.1073 |

**D. Confounder check — correlation of the first two principal components with slide quality metrics**

| Encoder | metric | r with PC1 | r with PC2 | median in FP | median in TN |
|---|---|---|---|---|---|
| TITAN (patch) | tissue_frac | -0.187 | -0.105 | 0.405 | 0.451 |
| TITAN (patch) | mean_gray | -0.799 | 0.109 | 242.5 | 245.1 |
| TITAN (patch) | tissue_mean_G | -0.841 | 0.096 | 234.5 | 232.9 |
| TITAN (patch) | slide_area_mm2 | -0.341 | 0.23 | 246.8 | 260.4 |
| Virchow2 | tissue_frac | 0.242 | -0.152 | 0.405 | 0.451 |
| Virchow2 | mean_gray | 0.843 | 0.026 | 242.5 | 245.1 |
| Virchow2 | tissue_mean_G | 0.897 | 0.03 | 234.5 | 232.9 |
| Virchow2 | slide_area_mm2 | 0.497 | 0.262 | 246.8 | 260.4 |
| GigaPath-256 | tissue_frac | -0.231 | -0.115 | 0.405 | 0.451 |
| GigaPath-256 | mean_gray | -0.842 | 0.013 | 242.5 | 245.1 |
| GigaPath-256 | tissue_mean_G | -0.883 | 0.012 | 234.5 | 232.9 |
| GigaPath-256 | slide_area_mm2 | -0.447 | 0.278 | 246.8 | 260.4 |
| Phikon-v2 | tissue_frac | 0.217 | -0.222 | 0.405 | 0.451 |
| Phikon-v2 | mean_gray | -0.903 | 0.139 | 242.5 | 245.1 |
| Phikon-v2 | tissue_mean_G | -0.937 | 0.181 | 234.5 | 232.9 |
| Phikon-v2 | slide_area_mm2 | -0.044 | 0.206 | 246.8 | 260.4 |
| H-Optimus-0 | tissue_frac | -0.253 | 0.011 | 0.405 | 0.451 |
| H-Optimus-0 | mean_gray | -0.84 | 0.133 | 242.5 | 245.1 |
| H-Optimus-0 | tissue_mean_G | -0.885 | 0.163 | 234.5 | 232.9 |
| H-Optimus-0 | slide_area_mm2 | -0.473 | 0.173 | 246.8 | 260.4 |
| GigaPath | tissue_frac | -0.249 | -0.115 | 0.405 | 0.451 |
| GigaPath | mean_gray | -0.848 | -0.007 | 242.5 | 245.1 |
| GigaPath | tissue_mean_G | -0.891 | -0.009 | 234.5 | 232.9 |
| GigaPath | slide_area_mm2 | -0.475 | 0.269 | 246.8 | 260.4 |
| Hibou-L | tissue_frac | 0.233 | 0.072 | 0.405 | 0.451 |
| Hibou-L | mean_gray | 0.868 | -0.032 | 242.5 | 245.1 |
| Hibou-L | tissue_mean_G | 0.911 | -0.061 | 234.5 | 232.9 |
| Hibou-L | slide_area_mm2 | 0.492 | 0.07 | 246.8 | 260.4 |
| UNI2-h | tissue_frac | -0.254 | -0.122 | 0.405 | 0.451 |
| UNI2-h | mean_gray | -0.838 | -0.044 | 242.5 | 245.1 |
| UNI2-h | tissue_mean_G | -0.879 | -0.065 | 234.5 | 232.9 |
| UNI2-h | slide_area_mm2 | -0.497 | 0.034 | 246.8 | 260.4 |

Footnotes

TP, true positive; TN, true negative; FP, false positive; FN, false negative; LOO, leave-one-case-out; IQR, interquartile range; PC, principal component; AUC, area under the receiver operating characteristic curve.

### Supplementary Table S5. Predicted multiplex immunofluorescence (virtual mIF) by fluorescence and histopathological group

| Rank | Channel | Statistic | TP | TN | FP | FN | FP − TN | permutation p |
|---|---|---|---|---|---|---|---|---|
| 1 | Cy5 | mean | 0.0084 | 0.0058 | 0.0023 | 0.016 | -0.0035 | 0.1063 |
| 2 | CD3 | frac | 0.0052 | 0.0049 | 0.0018 | 0.013 | -0.0031 | 0.1066 |
| 3 | PD-L1 | mean | 0.0005 | 0.0004 | 0.0002 | 0.0003 | -0.0003 | 0.146 |
| 4 | CK | frac | 0.0024 | 0.002 | 0.0007 | 0.0043 | -0.0013 | 0.2269 |
| 5 | **CD11c** | frac | 0.0021 | 0.001 | 0.0004 | 0.0028 | -0.0006 | 0.2389 |
| 6 | CK | mean | 0.0036 | 0.0034 | 0.0013 | 0.0078 | -0.0021 | 0.2479 |
| 7 | Transgelin | mean | 0.0006 | 0.0004 | 0.0001 | 0.0009 | -0.0003 | 0.2502 |
| 8 | PD-L1 | frac | 0.0073 | 0.0042 | 0.0021 | 0.0109 | -0.002 | 0.2562 |
| 9 | CD20 | frac | 0.001 | 0.0012 | 0.0007 | 0.0012 | -0.0005 | 0.2609 |
| 10 | TRITC | mean | 0.0024 | 0.0017 | 0.0011 | 0.0027 | -0.0006 | 0.2699 |
| 11 | CD4 | mean | 0.0039 | 0.0025 | 0.0011 | 0.0062 | -0.0013 | 0.2752 |
| 12 | CD8 | mean | 0.0043 | 0.0023 | 0.0013 | 0.0045 | -0.001 | 0.2882 |
| 13 | Actin | mean | 0.0045 | 0.0032 | 0.0016 | 0.0066 | -0.0015 | 0.3356 |
| 14 | Cy5 | frac | 0.0062 | 0.0028 | 0.0018 | 0.0061 | -0.001 | 0.3835 |
| 15 | **CD14** | frac | 0.0066 | 0.0039 | 0.002 | 0.0122 | -0.002 | 0.3972 |
| 16 | **CD14** | mean | 0.0105 | 0.0048 | 0.003 | 0.0142 | -0.0018 | 0.4079 |
| 17 | **CD16** | frac | 0.0017 | 0.0006 | 0.0003 | 0.0006 | -0.0003 | 0.4399 |
| 18 | CD34 | mean | 0.0012 | 0.0004 | 0.0003 | 0.0006 | -0.0002 | 0.4399 |
| 19 | TRITC | frac | 0.0063 | 0.0029 | 0.0018 | 0.0078 | -0.0011 | 0.4832 |
| 20 | CD3 | mean | 0.0034 | 0.0019 | 0.0008 | 0.0037 | -0.0011 | 0.4845 |
| 21 | Transgelin | frac | 0.0004 | 0.0003 | 0.0002 | 0.0004 | -0.0002 | 0.5065 |
| 22 | PHH3 | mean | 0.0004 | 0.0003 | 0.0001 | 0.0003 | -0.0002 | 0.5072 |
| 23 | T-bet | mean | 0.0003 | 0.0001 | 0.0001 | 0.0002 | -0.0001 | 0.5205 |
| 24 | T-bet | frac | 0.0029 | 0.0013 | 0.0009 | 0.0016 | -0.0003 | 0.5232 |
| 25 | PD-1 | mean | 0.0007 | 0.0003 | 0.0001 | 0.0004 | -0.0001 | 0.5285 |
| 26 | PD-1 | frac | 0.0022 | 0.0008 | 0.0006 | 0.0012 | -0.0002 | 0.5435 |
| 27 | Tryptase | frac | 0.0002 | 0.0001 | 0.0001 | 0.0001 | -0.0001 | 0.5475 |
| 28 | Tryptase | mean | 0.0016 | 0.0009 | 0.0006 | 0.0005 | -0.0003 | 0.5871 |
| 29 | Ki67 | mean | 0.0079 | 0.0017 | 0.0024 | 0.002 | 0.0007 | 0.6081 |
| 30 | CD8 | frac | 0.0001 | 0 | 0 | 0 | 0 | 0.6501 |
| 31 | CD20 | mean | 0.0101 | 0.0066 | 0.0049 | 0.0123 | -0.0016 | 0.6654 |
| 32 | CD138 | mean | 0.002 | 0.0006 | 0.0004 | 0.0008 | -0.0002 | 0.6741 |
| 33 | CD34 | frac | 0.0072 | 0.004 | 0.0029 | 0.0093 | -0.0011 | 0.6904 |
| 34 | Actin | frac | 0.0479 | 0.0117 | 0.0138 | 0.0333 | 0.0022 | 0.7014 |
| 35 | **CD16** | mean | 0.0009 | 0.0002 | 0.0002 | 0.0006 | 0 | 0.7154 |
| 36 | DAPI | frac | 0.0159 | 0.008 | 0.007 | 0.0183 | -0.0009 | 0.7408 |

| | | | | | | | | |
|---|---|---|---|---|---|---|---|---|
| 37 | CD4 | frac | 0.0123 | 0.0032 | 0.0034 | 0.0044 | 0.0002 | 0.7541 |
| 38 | **CD68** | frac | 0.0024 | 0.0006 | 0.0005 | 0.0015 | -0.0001 | 0.8221 |
| 39 | PHH3 | frac | 0.0324 | 0.0105 | 0.0097 | 0.0334 | -0.0009 | 0.8317 |
| 40 | **CD68** | mean | 0.052 | 0.0134 | 0.0152 | 0.038 | 0.0018 | 0.8371 |
| 41 | **CD11c** | mean | 0.0366 | 0.0112 | 0.0108 | 0.0358 | -0.0004 | 1.0 |
| 42 | CD138 | frac | 0.0421 | 0.0131 | 0.0126 | 0.0303 | -0.0006 | 1.0 |
| 43 | Caspase3 | frac | 0 | 0 | 0 | 0 | 0 | 1.0 |
| 44 | Caspase3 | mean | 0.0009 | 0.0002 | 0.0002 | 0.0005 | 0 | 1.0 |
| 45 | DAPI | mean | 0.0429 | 0.0133 | 0.0129 | 0.0316 | -0.0004 | 1.0 |
| 46 | Ki67 | frac | 0 | 0 | 0 | 0 | 0 | 1.0 |

Footnotes

mIF, multiplex immunofluorescence.

**Supplementary Table S6. Specimen-level fluorescence–pathology cross-tabulation (analysis set, n = 63)**

| # | Case | Specimen | Specimen name (as reported) | Fluorescence | Pathology (verbatim group) | Reference call | Classification |
|---|---|---|---|---|---|---|---|
| 1 | C01 | 5 | left temporal tumor (strong fluorescence) | Fluorescent | 2[frozen 1]/3[frozen 2]/4[frozen 3]/5[strong fluorescence]/6[weak fluorescence] | scattered tumor cells with post-treatment change → tumor | TP |
| 2 | C01 | 6 | left temporal tumor (weak fluorescence) | Fluorescent | same as above | scattered tumor cells with post-treatment change → tumor | TP |
| 3 | C02 | 2 | left temporal tumor (strong fluorescence) | Fluorescent | G1: post-frozen paraffin 1/2; 1, 2, 10, 11, 26, 27, 28, 31 | high-grade glioma → tumor | TP |
| 4 | C02 | 11 | left temporal tumor (5) (strong fluorescence) | Fluorescent | G1 | high-grade glioma → tumor | TP |
| 5 | C02 | 26 | periventricular fluorescent tissue 2 | Fluorescent | G1 | high-grade glioma → tumor | TP |
| 6 | C02 | 27 | periventricular fluorescent tissue 3 | Fluorescent | G1 | high-grade glioma → tumor | TP |
| 7 | C02 | 28 | periventricular tumor 2 (fluorescence) | Fluorescent | G1 | high-grade glioma → tumor | TP |
| 8 | C02 | 31 | left parieto-occipital cavity wall (weak fluorescence) | Fluorescent | G1 | high-grade glioma → tumor | TP |
| 9 | C02 | 6 | left temporal tumor (2) (strong fluorescence) | Fluorescent | G2: 3, 4, 5, 6, 7, 8, 9, 12–16, 17, 18, 19, 20–24, 29, 30 | peritumoral tissue → non-tumor | FP |
| 10 | C02 | 7 | left temporal tumor (4) (strong fluorescence) | Fluorescent | G2 | peritumoral tissue → non-tumor | FP |
| 11 | C02 | 9 | periventricular tissue (fluorescence) | Fluorescent | G2 | peritumoral tissue → non-tumor | FP |
| 12 | C02 | 18 | left temporal cavity, fluorescent tissue | Fluorescent | G2 | peritumoral tissue → non-tumor | FP |
| 13 | C02 | 19 | left temporal, intraoperative (strong fluorescence) | Fluorescent | G2 | peritumoral tissue → non-tumor | FP |
| 14 | C02 | 29 | left parieto-occipital cavity (strong fluorescence) | Fluorescent | G2 | peritumoral tissue → non-tumor | FP |
| 15 | C02 | 10 | periventricular tumor (no fluorescence) | Non-fluorescent | G1 | high-grade glioma → tumor | FN |
| 16 | C02 | 3 | left temporal tumor (no fluorescence) | Non-fluorescent | G2 | peritumoral tissue → non-tumor | TN |

| | | | | | | | |
|---|---|---|---|---|---|---|---|
| 17 | C02 | 4 | left temporal tissue (1) (no fluorescence) | Non-fluorescent | G2 | peritumoral tissue → non-tumor | TN |
| 18 | C02 | 5 | left temporal tissue (3) (no fluorescence) | Non-fluorescent | G2 | peritumoral tissue → non-tumor | TN |
| 19 | C02 | 8 | periventricular tissue (no fluorescence) | Non-fluorescent | G2 | peritumoral tissue → non-tumor | TN |
| 20 | C02 | 20 | left temporal, intraoperative, no fluorescence 1 | Non-fluorescent | G2 | peritumoral tissue → non-tumor | TN |
| 21 | C02 | 21 | left temporal, intraoperative, no fluorescence 2 | Non-fluorescent | G2 | peritumoral tissue → non-tumor | TN |
| 22 | C02 | 22 | left temporal, intraoperative, no fluorescence 3 | Non-fluorescent | G2 | peritumoral tissue → non-tumor | TN |
| 23 | C02 | 23 | left temporal, intraoperative, no fluorescence 4 | Non-fluorescent | G2 | peritumoral tissue → non-tumor | TN |
| 24 | C02 | 24 | left parieto-occipital, intraoperative, no fluorescence | Non-fluorescent | G2 | peritumoral tissue → non-tumor | TN |
| 25 | C03 | 2 | right parietal tumor 1 (strong fluorescence, navigation yellow) | Fluorescent | findings consistent with high-grade glioma (2, 3, 4, 5, 6, 7, 8, 9, 10, 11 and post-frozen paraffin) | high-grade glioma → tumor | TP |
| 26 | C03 | 3 | right parietal tumor 2 (strong fluorescence, navigation red) | Fluorescent | same as above | high-grade glioma → tumor | TP |
| 27 | C03 | 4 | cavity (fluorescence) | Fluorescent | same as above | high-grade glioma → tumor | TP |
| 28 | C03 | 5 | cavity (fluorescence 2) | Fluorescent | same as above | high-grade glioma → tumor | TP |
| 29 | C03 | 6 | cavity (no fluorescence) | Non-fluorescent | same as above | **non-tumor (reclassified on per-specimen review)** | **TN** |
| 30 | C03 | 7 | ventricular wall (fluorescence) | Fluorescent | same as above | high-grade glioma → tumor | TP |
| 31 | C03 | 8 | corpus callosum tumor (fluorescence) | Fluorescent | same as above | high-grade glioma → tumor | TP |
| 32 | C03 | 9 | right parietal tumor, bulk 1 (no fluorescence) | Non-fluorescent | same as above | **non-tumor (reclassified on per-specimen review)** | **TN** |

| | | | | | | | |
|---|---|---|---|---|---|---|---|
| 33 | C03 | 10 | right parietal tumor, bulk 1 (fluorescence) | Fluorescent | same as above | high-grade glioma → tumor | TP |
| 34 | C03 | 11 | right parietal tumor, bulk 2 (fluorescence) | Fluorescent | same as above | high-grade glioma → tumor | TP |
| 35 | C04 | 2 | frontal tumor 1 (fluorescence A) | Fluorescent | G2: 2, 3, 4, 6, 8, 9, 12, 14, 15, 16 | IDH-wildtype glioma, high grade favored → tumor | TP |
| 36 | C04 | 3 | left frontal tumor 2 (fluorescence B) | Fluorescent | G2 | same as above → tumor | TP |
| 37 | C04 | 4 | left frontal tumor 3 (fluorescence C) | Fluorescent | G2 | same as above → tumor | TP |
| 38 | C04 | 6 | posterior margin, intraoperative fluorescence | Fluorescent | G2 | same as above → tumor | TP |
| 39 | C04 | 8 | medial margin, intraoperative strong fluorescence | Fluorescent | G2 | same as above → tumor | TP |
| 40 | C04 | 9 | anterior margin, intraoperative fluorescence | Fluorescent | G2 | same as above → tumor | TP |
| 41 | C04 | 12 | anterior margin, intraoperative fluorescence 2 | Fluorescent | G2 | same as above → tumor | TP |
| 42 | C04 | 14 | tumor body, fluorescence 1 | Fluorescent | G2 | same as above → tumor | TP |
| 43 | C04 | 15 | tumor body, fluorescence 2 | Fluorescent | G2 | same as above → tumor | TP |
| 44 | C04 | 5 | left frontal lesion (no fluorescence D) | Non-fluorescent | G3: post-frozen paraffin, 5, 7, 10, 11, 13 | glioma tissue identified → tumor | FN |
| 45 | C04 | 7 | posterior margin, intraoperative, no fluorescence | Non-fluorescent | G3 | glioma tissue identified → tumor | FN |
| 46 | C04 | 10 | anterior margin, intraoperative, no fluorescence | Non-fluorescent | G3 | glioma tissue identified → tumor | FN |
| 47 | C04 | 11 | anterior margin, intraoperative, no fluorescence 2 | Non-fluorescent | G3 | glioma tissue identified → tumor | FN |
| 48 | C04 | 13 | cavity floor, fluorescence | Fluorescent | G3 | glioma tissue identified → tumor | TP |
| 49 | C05 | 1 | left frontal tumor surface, fluorescence 1 | Fluorescent | consistent with high-grade glioma (post-frozen paraffin, 1, 2, 3, 4, 6, 8) | high-grade glioma → tumor | TP |

| | | | | | | | |
|---|---|---|---|---|---|---|---|
| 50 | C05 | 2 | left frontal tumor, intraoperative fluorescence 2 | Fluorescent | same as above | high-grade glioma → tumor | TP |
| 51 | C05 | 3 | left frontal cavity, fluorescence | Fluorescent | same as above | high-grade glioma → tumor | TP |
| 52 | C05 | 4 | left frontal cavity, no fluorescence | Non-fluorescent | same as above | high-grade glioma → tumor | FN |
| 53 | C05 | 6 | left frontal cavity, no fluorescence 3 | Non-fluorescent | same as above | high-grade glioma → tumor | FN |
| 54 | C05 | 5 | left frontal cavity, no fluorescence 2 | Non-fluorescent | listed separately: interpreted as reactive hyperplasia | non-tumor | TN |
| 55 | C06 | 1 | left frontal tumor, intraoperative fluorescence 1 | Fluorescent | G3: 1, 3, 4, 6, 9 | glioma favored → tumor | TP |
| 56 | C06 | 3 | left frontal, navigation fluorescence 1 | Fluorescent | G3 | glioma favored → tumor | TP |
| 57 | C06 | 4 | left frontal cavity, fluorescence 3 | Fluorescent | G3 | glioma favored → tumor | TP |
| 58 | C06 | 6 | left frontal cavity, fluorescence 5 | Fluorescent | G3 | glioma favored → tumor | TP |
| 59 | C06 | 2 | left frontal tumor, intraoperative fluorescence 2 | Fluorescent | G2: post-frozen paraffin 2, 2, 5 | glioma with post-treatment change → tumor | TP |
| 60 | C06 | 5 | left frontal cavity, fluorescence 4 | Fluorescent | G2 | glioma with post-treatment change → tumor | TP |
| 61 | C06 | 7 | left frontal cavity, no fluorescence 1 | Non-fluorescent | G1: post-frozen paraffin 1, 7, 8 | reactive hyperplasia → non-tumor | TN |
| 62 | C06 | 8 | left frontal cavity, fluorescence 6 | Fluorescent | G1 | reactive hyperplasia → non-tumor | FP |
| 63 | C06 | 9 | left frontal cavity, no fluorescence 2 | Non-fluorescent | G3 | glioma favored → tumor | FN |

**Total: 35 true positive, 7 false positive, 8 false negative and 13 true negative.**

Footnotes

TP, true positive; FP, false positive; FN, false negative; TN, true negative.

**Supplementary Table S7. Video-derived intraoperative observation times**

| Case | Video fragments | White light, min | Blue light, no fluorescence, min | Blue light with tumor fluorescence, min | Blue light, min | Fluorescence segments |
|---|---|---|---|---|---|---|
| C01 | 25 | 158.4 | 9.1 | 15.7 | 24.8 | 53 |
| C02 | 36 | 208.0 | 18.5 | 33.1 | 51.6 | 130 |
| C03 | 20 | 106.8 | 17.1 | 21.2 | 38.3 | 109 |
| C04 | 31 | 155.5 | 6.5 | 29.0 | 35.5 | 102 |
| C05 | — | Not recorded | Not recorded | Not recorded | Not recorded | — |
| C06 | 6 | 48.7 | 0.3 | 1.0 | 1.3 | 7 |
| **Total** | **118** | **677.5** | **51.5** | **100.0** | **151.5** | **401** |

### Supplementary Table S8. Perioperative safety

| Case | Intraoperative blood loss | Transfusion | Intraoperative neurophysiology | Postoperative complications | Light-avoidance instruction |
|---|---|---|---|---|---|
| C01 | 400 mL | None | **SEP/MEP stable** | Uneventful recovery | Environmental light avoidance; avoid examining the pupil |
| C02 | 400 mL | FFP 200 mL | — | **CNS infection** | Environmental light avoidance; avoid examining the pupil |
| C03 | 400ml | None | — | Uneventful recovery | Environmental light avoidance; avoid examining the pupil |
| C04 | Not reported | None | **SEP/MEP stable** | Uneventful recovery | Environmental light avoidance; avoid examining the pupil |
| C05 | 200 mL | None | **SEP/MEP stable** | Uneventful recovery | Environmental light avoidance; avoid examining the pupil |
| C06 | 200 mL | None | **SEP/MEP stable** | Uneventful recovery | Environmental light avoidance; avoid examining the pupil |

Footnotes

FFP, fresh frozen plasma; SEP, somatosensory evoked potential; MEP, motor evoked potential; CNS, central nervous system.

### Supplementary Table S9. Reported diagnostic performance of fluorescence-guided resection in glioma

| Study | Patients/Specimens | Level | Agent | Sensitivity | Specificity | Notes |
|---|---|---|---|---|---|---|
| Hauser et al, 2016[27] | 12 patients / 117 specimens | specimen | 5-ALA | 81% | 43% | Only 7 non-neoplastic specimens |
| Aibar-Durán et al, 2025[28] | 72 patients / 301 biopsies | specimen | 5-ALA (any positive or weak) | 84.9% | 57.8% | AUC 0.80 |
| Dablouk et al, 2024[29] | 28 patients / 83 samples | specimen | 5-ALA fiber probe | 73.3% | 100% | No false positives; NPV 28.6% |
| Valdés et al, 2015[30] | 12 patients / 73 specimens | specimen | 5-ALA (visual fluorescence) | 21% | 81% | Low-grade glioma |
| Watts et al, 2023[31] | 81 fluorescent / 9 non-fluorescent patients | patient | 5-ALA (GALA-BIDD) | 100% | 88.9% | Target condition: high-grade lesion |
| Ling et al, 2024[32] | HGG subgroup | specimen | fluorescein 1 mg/kg | 93.5% | 82.6% | — |
| Chen et al, 2019[33] | 49 patients / 98 biopsies | specimen | fluorescein 5 mg/kg | 91.7% | 90.0% | Retrospective |
| **Present study** | **6 patients / 63 specimens** | specimen | HpD (Hiporfin) | 75.1% | 60.6% | AUC 0.739 |

Footnotes

5-ALA, 5-aminolevulinic acid; HpD, hematoporphyrin derivative; HGG, high-grade glioma; NPV, negative predictive value; AUC, area under the receiver operating characteristic curve; FGR, fluorescence-guided resection.

### Supplementary figure legends

#### Supplementary Figure S1

**Supplementary Figure S1.** Two-level structure of the bivariate random-effects model used to pool diagnostic accuracy (6 patients, 63 specimens). **A**, Level 1 (within-patient sampling): each patient contributes a 2 × 2 table of true-positive (TP), false-positive (FP), false-negative (FN) and true-negative (TN) specimen counts, modeled with binomial likelihoods; blue cells denote concordant and orange cells discordant classifications, and the number of specimens examined per patient ranged from 2 to 22. **B**, Level 2 (between-patient distribution): patient-specific pairs of logit sensitivity and logit false-positive rate are assumed to be drawn from a bivariate normal distribution with mean $\mu$ and covariance $\Sigma$; point size is proportional to the number of specimens examined per patient, the filled diamond is the pooled estimate and the shaded ellipse its 95% confidence region; $\sigma$_Se and $\sigma$_FPR are the between-patient standard deviations, both estimated at essentially zero in this cohort ($I^2 = 10.7\%$). **C**, Interpretation of the between-patient correlation $\rho$: movement along the summary ROC curve (green) is a shift in diagnostic threshold, with sensitivity and false-positive rate rising together ($\rho > 0$), whereas movement away from the curve towards the upper left (purple) represents better overall accuracy, with sensitivity rising at a fixed false-positive rate ($\rho < 0$). In this cohort $\rho$ could not be estimated reliably, but the pooled estimates were insensitive to the assumption made about $\rho$. FPR = false-positive rate; ROC = receiver operating characteristic.

#### Supplementary Figure S2

**Supplementary Figure S2.** Serial MRI of the two lesions resected through a single craniotomy in the patient who underwent resection of two lesions. **A**, the newly diagnosed left temporal lesion; **B**, the recurrent left parieto-occipital lesion. In **A** and **B** the columns show the same lesion before surgery and at four time points after surgery (1 week, and 3, 6 and 12 months), and the rows show six sequences: T1-weighted, T2-weighted and T2-weighted fluid-attenuated inversion recovery images, followed by contrast-enhanced T1-weighted images in the axial, coronal and sagittal planes. Images were cropped to a constant physical field of view (190 × 190 mm for axial images, 175 × 205 mm for coronal and sagittal images, centered on the brain) and scaled proportionally, so that the two lesions and the successive time points are displayed at a directly comparable scale; the anatomical level shown was selected at each time point to include the lesion and is therefore similar, but not identical, across the columns. Each of **A** and **B** comprises 30 images. MRI = magnetic resonance imaging.

#### Supplementary Figure S3

**Supplementary Figure S3.** Paired preoperative and postoperative axial T1-weighted contrast-enhanced images for all six patients, three levels per patient (left, preoperative; right, postoperative). Each pair is separated from the next by a wider gap. Panels are shown in radiological orientation (top, anterior; right, patient's left) on a common 160-mm field. Postoperative images were acquired on day 1–2 after surgery (day 7 in C02); the postoperative panel of C01 is the same-day CT in brain window, as no postoperative MRI was obtained in that patient. Relative windowing was applied uniformly across panels (each volume mapped from its own 2nd–98th brain-intensity percentiles), so intensities are not directly comparable between modalities. Illustrative only; extent of resection was not verified volumetrically (see Discussion, limitations).

#### Supplementary Figure S4

**Supplementary Figure S4.** Morphological comparison of HE-stained slides according to intraoperative fluorescence status and histopathological diagnosis using eight frozen pathology-encoder configurations applied to HE-stained slides (42 true-positive, 15 true-negative, 12 false-negative and 7 false-positive HE-stained slides; 76 slides in total). Seven distinct tile-level encoders were used (Virchow2, Phikon-v2, H-Optimus-0, Hibou-L, UNI2-h, GigaPath and the TITAN/CONCH v1.5 patch encoder); GigaPath was also evaluated at its native 256-px field size and is therefore listed twice (GigaPath and GigaPath-256). The descriptor of a slide was the L2-normalized mean of its tile feature vectors, so no slide-level spatial aggregation was used in this figure. **A**, For each encoder, the cosine distance between the centroid of the false-positive specimens and those of the true-positive (red), false-negative (green) and true-negative (blue) specimens. Symbols are the observed distances, horizontal lines the range obtained by leave-one-case-out resampling, and p values two-sided permutation tests (2,000 permutations). False-positive specimens were significantly distant from true-positive specimens for every encoder ($p = 0.001$–$0.038$; leave-one-case-out ranges 0.021–0.111, none including zero) and from false-negative specimens in seven of eight encoders (distances 0.0149–

0.0726; p = 0.0155–0.053; H-Optimus-0, p = 0.053; leave-one-case-out ranges 0.0176–0.0919, none including zero), whereas no encoder separated false-positive from true-negative specimens (p = 0.234–0.430). The distance from false positives thus increased monotonically from true-negative to false-negative to true-positive specimens. **B**, Tumor-similarity difference (cosine similarity to the true-positive centroid minus cosine similarity to the true-negative centroid; values above zero indicate greater morphological similarity to tumor than to non-neoplastic tissue) for each specimen, ordered consistently across all eight encoders. True-positive specimens had positive values, false-negative specimens were intermediate and close to zero, and false-positive and true-negative specimens had negative values. Values are group medians with interquartile ranges. Tile-level features were extracted at 512 × 512 px and 0.5 µm/px; all encoders shared the same tile coordinates. Four of the eight configurations (Phikon-v2, UNI2-h, Virchow2 and TITAN) are released for non-commercial research use only. Nominal p values are uncorrected for multiple comparisons; Supplementary Table S4 gives all comparisons, dimensions, revisions and license terms, and Supplementary Table S3 gives the analysis set. HE = hematoxylin and eosin.

### Supplementary Figure S5

**Supplementary Figure S5.** Intraoperative frames exported during the analysis but not displayed in Figure 4. **Left column**, tumor fluorescence: frame pair #12, which duplicates the surgical field shown by the #13 pair in Figure 4A (upper image, white light; lower image, blue light). **Right column**, cavity fluorescence: frame pair #34, left parieto-occipital field (upper image, white light; lower image, blue light).
