## Supplementary material for "Hiporfin (HpD) for fluorescence-guided resection of recurrent glioblastoma: diagnostic accuracy and feasibility of an integrated photodynamic strategy": STRAD 2015 Checklist

### STARD 2015 checklist — Hiporfin (hematoporphyrin derivative) for fluorescence-guided resection of recurrent glioblastoma

**Manuscript:** *Hiporfin (HpD) for fluorescence-guided resection of recurrent glioblastoma: diagnostic accuracy and feasibility of an integrated photodynamic strategy*

**Target journal:** *Neuro-Oncology* (submission track; the manuscript is also formatted to the Journal of Neurosurgery clinical-article limits) | **Article type for reporting purposes:** diagnostic accuracy study embedded in a prospective single-arm feasibility series

**Reporting guideline:** STARD 2015[1] (30 items → 34 rows) | **Checklist completed:** 2026-09-21 | **Last revised: 2026-10-04** | **Filled by:** Zhenqiang He

**Corresponding author / signature:** Yonggao Mou, Xiangheng Zhang

***Scope.*** *This checklist covers the diagnostic-accuracy component only (index test = intraoperative fluorescence assessment; reference standard = histopathology of the resected specimen). Workflow feasibility and perioperative safety (secondary endpoints) are reported descriptively and are outside the scope of the STARD items, except item 25 (adverse events).*

#### Checklist

| # | STARD 2015 item (verbatim) | Location | Status, content and outstanding action |
|---|---|---|---|
| **1** | Identification as a study of diagnostic accuracy using at least one measure of accuracy (such as sensitivity, specificity, predictive values, or AUC) | Title page; Abstract | ✓ **In place.** The title carries "diagnostic accuracy"; sensitivity, specificity, PPV, NPV and AUC are reported in the text and the abstract, and "diagnostic accuracy" is a keyword. |
| **2** | Structured summary of study design, methods, results, and conclusions (for specific guidance, see STARD for Abstracts) | Abstract | ✓ **In place. 228 words** (Neuro-Oncology limit 250), structured **Background / Methods / Results / Conclusions**, no citations, five keywords. Contains: prospective single-arm design and center; enrolment (6 patients / 7 resection fields / 63 specimens); index test (intraoperative fluorescence under blue-light excitation) and reference standard (histopathology); pooled sensitivity 0.751 (0.600–0.858) and specificity 0.606 (0.407–0.775) with 95% CIs and AUC 0.739; the concentration of false positives in peritumoral tissue; and the statement that no efficacy claim is supported. |
| **3** | Scientific and clinical background, including the intended use and clinical role of the index test | Introduction ¶1–¶5 | ✓ **In place.** The background covers the limits of reoperation for recurrent glioblastoma, the EANO/NCCN positioning, the difficulty of distinguishing residual tumor from radiation necrosis and gliotic scar under white light, the established role and limitations of 5-ALA (including its false-positive behavior in reactive tissue), the precedent of 5-ALA + intraoperative PDT, and the diagnostic–therapeutic duality of porphyrins with the 1987 HpD PDT experience. The **intended use** and **clinical role** of the index test are stated in the aim sentence of Introduction ¶5: Hiporfin fluorescence is described as **an adjunct for intraoperative margin guidance during repeat resection**. The consequence for practice — that absent fluorescence does not license stopping the resection — is stated in Discussion (principal findings): "fluorescence alone cannot declare a margin clear" (item 27). |

| | | | |
|---|---|---|---|
| **4** | Study objectives and hypotheses | Introduction ¶5 | ✓ **In place.** Two-step objective stated: ① quantify the diagnostic performance of Hiporfin fluorescence at the specimen level against histopathology; ② establish whether the one-drug / one-dose / one-wavelength fluorescence-guided resection + PDT workflow is feasible. **No formal statistical hypothesis** (exploratory feasibility positioning), which Methods states. |
| **5** | Whether data collection was planned before the index test and reference standard were performed (prospective study) or after (retrospective study) | Methods §Study design (¶1) | ✓ **In place.** "This prospective, single-arm case series was performed at one tertiary neurosurgical center…". The same paragraph discloses the registration timing (NCT07832448 first posted after enrolment was complete) and states that this analysis was not a prespecified endpoint of the registration record. |
| **6** | Eligibility criteria | Methods §Patients | ✓ **In place.** Eligibility criteria are stated as prespecified in the registered protocol (ClinicalTrials.gov **NCT07832448**): adults 18–65 years with histopathologically confirmed glioblastoma recurring or progressing after standard radiotherapy and chemotherapy; KPS ≥ 70; measurable disease on imaging; ≥ 3 months since brain radiotherapy; lesion considered resectable at reoperation. Principal exclusions: KPS < 70, prior or concurrent malignancy, contraindications to surgery or MRI, porphyria or soy allergy, severe renal or hepatic impairment, pregnancy. The full registered list (16 inclusion, 18 exclusion) is public in the registration record. |
| **7** | On what basis potentially eligible participants were identified (such as symptoms, results from previous tests, inclusion in registry) | Methods §Patients | ✓ **In place.** "Potentially eligible patients were identified prospectively: patients with a previously histopathologically confirmed glioblastoma who presented at our institution with brain MRI findings suggestive of local recurrence were referred to the study team for screening." The cohort definition (all patients in the institutional recurrent-glioblastoma programme undergoing Hiporfin fluorescence-guided resection plus intraoperative PDT) is given in the same paragraph. |
| **8** | Where and when potentially eligible participants were identified (setting, location and dates) | Methods §Study design; Table 1 (per case) | ✓ **In place.** Single tertiary neurosurgical center; surgery performed September–October 2024, with per-case month and year in Table 1. The full name of the center is completed on the title page together with the author list. |
| **9** | Whether participants formed a consecutive, random or convenience series | Methods §Patients | ✓ **In place.** Patients were enrolled **consecutively** in a prospective institutional protocol of microsurgical resection combined with Hiporfin PDT for recurrent glioblastoma. |
| **10a** | Index test, in sufficient detail to allow replication | Methods §Procedures (dosing; resection and fluorescence assessment); §Statistical analysis | ✓ **In place.** Written: (i) single intravenous Hiporfin **5 mg/kg** (230–400 mg administered; Table 1), skin test before infusion, no concurrent photosensitising agent; (ii) per-case timing — **dosing → incision** |

| | | | |
|---|---|---|---|
| | | | **40.0–46.3 h** (median 45.6 h; Table 1) and **dosing → PDT illumination 46.9–53.2 h** (median 52.6 h), **6/6 within the protocol window of 40–72 h and 5/6 within the label window of 48–72 h** (C06, 46.9 h); (iii) resection under white light followed by blue-light excitation, fluorescence graded by the operating surgeon as strong / moderate / weak / absent and specimen labels assigned accordingly; (iv) fluorophore and light-discrimination rules; (v) **imaging platform** ZEISS Kinevo 900 in BLUE 400 mode (400–410 nm excitation / 620–710 nm detection), with the disclosure that this was derived from the operative video because it was not recorded in the surgical record. |
| **10b** | Reference standard, in sufficient detail to allow replication | Methods §Specimen sampling and reference standard; Results §Diagnostic accuracy; Table 2 footnotes; **Supplementary Table S3** | ✓ **In place.** Reference standard = the histopathological diagnosis of the corresponding specimen, issued as "the written histopathological report by pathologists specializing in neuro-oncology" (a routine diagnostic report, not a study-specific reading); per-specimen reports are listed verbatim in Supplementary Table S3. The tumor/non-tumor classification rule is written, as is the reclassification of two non-fluorescent C03 specimens as non-tumor on per-specimen review (Table 2, footnote). |
| **11** | Rationale for choosing the reference standard (if alternatives exist) | Methods §Specimen sampling and reference standard | ✓ **In place.** The rationale is the same-specimen design: each histopathological diagnosis is assigned to the same specimen that carried the fluorescence label, so the index test and the reference standard describe identical tissue; the alternatives used in previous quantitative studies (proliferation markers, cell density) are identified in the Introduction as not answering whether a specimen contains tumor. |
| **12a** | Definition of and rationale for test positivity cut-offs or result categories of the index test, distinguishing pre-specified from exploratory | Methods §Statistical analysis | ✓ **In place.** Strong, moderate and weak fluorescence were pooled as "positive" and absent fluorescence as "negative"; the pooling was prespecified, not post hoc; intensity is a subjective call and the numbers are small, so no continuous threshold exists and ROC threshold selection does not apply. |
| **12b** | Definition of and rationale for test positivity cut-offs or result categories of the reference standard, distinguishing pre-specified from exploratory | — | **N/A.** The reference standard is a categorical histopathological diagnosis (tumor / non-tumor) with no adjustable positivity threshold, so the threshold component does not apply. The classification rule is presented per specimen in the Table 2 footnotes and Supplementary Table S3. |
| **13a** | Whether clinical information and reference standard results were available to the performers/readers of the index test | Methods §Statistical analysis | ✓ **In place.** When reading fluorescence the operating surgeon knew the clinical history and previous imaging but did not know the histopathological result at that time — both consistent with how the index test was actually used. |
| **13b** | Whether clinical information and index test results | Methods §Statistical analysis; Discussion | ❌ **Not satisfied — assessors were not blinded; the** |

| | | | |
|---|---|---|---|
| | were available to the assessors of the reference standard | (limitations) | **reporting obligation is fulfilled.** Methods states that the histopathological assessors were not blinded to the intraoperative fluorescence status because the specimen labels carried the fluorescence description, and that this is reported as a potential source of bias (STARD 2015 item 13b is not satisfied); Discussion (limitations) repeats it. |
| **14** | Methods for estimating or comparing measures of diagnostic accuracy | Methods §Statistical analysis | ✓ **In place.** The patient is the unit of analysis; specimen-level crude pooling with Wilson 95% CIs (descriptive); Reitsma bivariate random-effects model (0.5 continuity correction, REML, $I^2$, SROC curve and AUC; partial AUC reported); when ρ reached a boundary a univariate random-effects model (ρ = 0) was used as a sensitivity analysis; software R 4.6.1 / mada 0.5.12 / metafor 5.0.1. |
| **15** | How indeterminate index test or reference standard results were handled | Methods §Statistical analysis; Results §Diagnostic accuracy | ✓ **In place.** Specimens without an explicit fluorescence annotation (frozen-section, uncut gross, unannotated peritumoral tissue) and dura specimens were excluded from the primary analysis; the two fluorescent-labeled dura specimens were included in a prespecified sensitivity analysis (n = 65). The per-category composition of the 24 excluded specimens is given in **Supplementary Table S2**. |
| **16** | How missing data on the index test and reference standard were handled | Methods §Statistical analysis; Results §Diagnostic accuracy | ✓ **In place.** Specimens with a missing fluorescence annotation were excluded and their composition is described; specificity was not estimable in the two patients without non-tumor specimens (C01, C04), which therefore do not enter the SROC analysis — stated in Results and in the Table 2 footnotes; zero cells were handled with a 0.5 continuity correction. |
| **17** | Any analyses of variability in diagnostic accuracy, distinguishing pre-specified from exploratory | Methods §Statistical analysis; Results §Diagnostic accuracy | ✓ **In place.** Leave-one-patient-out sensitivity analysis; the prespecified n = 65 sensitivity analysis including the two dura specimens; the comparison of the two resection fields in C02 (explicitly labeled descriptive/exploratory, with Fisher p = 1.00 and no quantitative claim); and the exploratory morphology comparison, labeled exploratory throughout. |
| **18** | Intended sample size and how it was determined | Methods §Study design (final sentence) | ✓ **In place.** The manuscript states that no a priori sample size calculation was performed and explains that the cohort comprises all patients completing the integrated workflow in the institutional recurrent-glioblastoma programme during the study period — internally consistent for an exploratory feasibility study. |
| **19** | Flow of participants, using a diagram | Figure 1B | ✓ **In place.** Figure 1B is the STARD flow diagram: 10 patients screened → 4 excluded (PDT without fluorescence-guided resection) → 6 patients (7 resection fields) → 87 specimens submitted → 24 excluded (8 frozen-section, 5 uncut gross, 5 dura, 2 peritumoral, 4 without fluorescence annotation) → 63 paired specimens analyzed. Figure 1A shows the single-operation workflow (5 mg/kg dosing → |

| | | | |
|---|---|---|---|
| | | | fluorescence-guided resection → 630 nm / 150 J/cm² cavity PDT). |
| **20** | Baseline demographic and clinical characteristics of participants | Table 1; Results §Patient characteristics and treatment | ✓ **In place.** Reported: sex, age, primary diagnosis, date of surgery (month and year), body weight, dose, dosing-to-incision interval, **MGMT promoter status** (molecular 4/6; immunohistochemistry-inferred 2/6), KPS, extent of resection, Ki-67 index and follow-up (now in Discussion). Weights 80/49/49/58/46/57 kg and KPS 80/90/70/100/80/90; dose–weight check 6/6 concordant. Extent of resection is surgeon-assessed and was **not** verified volumetrically (see Discussion, limitations). |
| **21a** | Distribution of severity of disease in those with the target condition | Results §Patient characteristics and treatment; Table 1 | ✓ **In place.** Results reports the Ki-67 index recorded in the diagnostic pathology reports (**5–60%** across the six patients; Table 1) as the measure of disease severity. All tumors were IDH-wildtype glioblastoma, WHO CNS grade 4. |
| **21b** | Distribution of alternative diagnoses in those without the target condition | Results §Diagnostic accuracy; Table 2 footnotes; **Supplementary Table S3** | ✓ **In place.** All seven false-positive specimens were reported histopathologically as **peritumoral tissue** (not tumor); the true-negative specimens were peritumoral tissue or post-treatment change. Listed per specimen (63 rows) in Supplementary Table S3. |
| **22** | Time interval and any clinical interventions between index test and reference standard | Methods §Specimen sampling (same specimen); §Procedures (dosing); Results §Diagnostic accuracy and §Feasibility and safety | ✓ **In place.** (i) The reference standard is the histopathological report of **the same specimen** (separate container labeled with site and fluorescence status), so no interval has to be bridged; (ii) the per-case dosing-to-incision interval is reported in Methods and Table 1 (**40.0–46.3 h**, median 45.6 h); (iii) PDT was delivered **after** sampling (630 nm, 150 J/cm²; 6 patients, 7 fields) — this alters the operative field but not the histopathological classification of specimens already removed. |
| **23** | Cross tabulation of the index test results (or their distribution) by the results of the reference standard | Table 2; **Supplementary Table S3**; Results §Diagnostic accuracy | ✓ **In place.** Pooled 2 × 2: **TP 35 / FP 7 / FN 8 / TN 13 (n = 63)**; per-patient 2 × 2 tables with sensitivity, specificity and 95% CIs in Table 2; the 63-row per-specimen fluorescence–pathology cross-tabulation in Supplementary Table S3. Exclusions are accounted for under items 15 and 16; no result was silently discarded. |
| **24** | Estimates of diagnostic accuracy and their precision (such as 95% confidence intervals) | Results §Diagnostic accuracy; Table 2; Figure 2A and 2B | ✓ **In place.** Bivariate pooled **sensitivity 0.751 (0.600–0.858)**, **specificity 0.606 (0.407–0.775)**, **AUC 0.739** (partial AUC 0.685), $I^2$ 10.7%; crude specimen-level sensitivity 0.814 (0.674–0.903), specificity 0.650 (0.433–0.819), PPV 0.833 (0.694–0.917), NPV 0.619 (0.409–0.792), accuracy 0.762 (0.644–0.850); leave-one-patient-out ranges reported. The video-derived durations do not enter the accuracy estimates; they are reported descriptively in **Supplementary Table S7** (151.5 and 100.0 min). |
| **25** | Any adverse events from performing the index test | Results §Feasibility and safety; **Supplementary** | ✓ **In place.** The index test (intraoperative |

| | | | |
|---|---|---|---|
| | or the reference standard | **Table S8** | fluorescence assessment) and the reference standard (histopathological examination) are not separate procedures and caused no adverse events. Safety is reported for the operation as a whole: estimated blood loss **mean 300 mL (range 200–400 mL; all six patients recorded)**, 200 mL fresh frozen plasma in C02, stable somatosensory/motor evoked potentials where monitored, no new postoperative neurological deficit, one CNS infection (C02), five uneventful courses. Cutaneous phototoxicity remains reported as "**not documented**" (absence of a record is not absence of an event); the Supplementary Table S8 footnote says so. |
| **26** | Study limitations, including sources of potential bias, statistical uncertainty, and generalisability | Discussion (limitations paragraph) | ✓ **In place.** Written: small single-arm series (6 patients / 7 fields / 63 specimens), hypothesis-generating, no efficacy claim; specificity resting on four patients; between-patient correlation and the binary surgeon call; C02's two lesions entering as one unit; **assessors not blinded** (item 13b); the exploratory morphology analysis with anti-conservative nominal p values (false positives from two patients only); **extent of resection not verified volumetrically** (from the operative note; postoperative contrast-enhanced MRI available in five of six); incomplete follow-up (two of six lost); photosensitivity assessed retrospectively; and that the analysis was **not a prespecified endpoint of a study registered after enrolment was complete**, so the results are a post hoc evaluation of prospectively collected material. |
| **27** | Implications for practice, including the intended use and clinical role of the index test | Discussion (principal findings; specificity paragraph; accessibility paragraph); Conclusions | ✓ **In place.** Explicit: a fluorescent region is very likely tumor (PPV 0.833) whereas a non-fluorescent region frequently still contains tumor (NPV 0.619) and **"fluorescence alone cannot declare a margin clear"**; the source of error is peritumoral tissue of the previously treated cavity, not the drug or the optics; Hiporfin is an available drug (licensed in China since 2001) while 5-ALA is not approved in mainland China; standardized prospective acquisition is required before thresholds can replace visual inspection. **The explicit "intended use / clinical role" sentence is the outstanding action under item 3.** |
| **28** | Registration number and name of registry | Methods (¶1); title page | ✓ **In place.** Registry **ClinicalTrials.gov**, number **NCT07832448** (https://clinicaltrials.gov/study/NCT07832448), first posted 2026-09-22. The registered study is the **parent series of 10 patients**; this report presents the **6 patients** who received both fluorescence-guided resection and PDT. Registration **post-dates completion of enrolment** (registered Start Date 2023-08-04) and the diagnostic-accuracy analysis |

| | | | |
|---|---|---|---|
| | | | was not a prespecified endpoint — disclosed in Methods ¶1 and Discussion (limitations). |
| **29** | Where the full study protocol can be accessed | Statements §6 Data availability | ✓ **In place.** The protocol is publicly posted on the registration platform (**ClinicalTrials.gov NCT07832448**); Statements §6 states that the study protocol is registered and publicly posted there and that the registration record will be submitted with the manuscript. |
| **30** | Sources of funding and other support; role of funders | Statements §1 Funding; §2 Competing interests | ✓ **In place.** Investigator-initiated, no industry sponsor: the sponsor-investigator in the registration is the corresponding author, and the authors designed and conducted the study independently. **Hiporfin was supplied by Chongqing Milelonge Biopharmaceutical Co., Ltd. (Chongqing, China), the manufacturer**, as non-financial support (drug supply) — declared in Statements §1 and §2, together with the authors' declaration that they have no financial relationships with the manufacturer (no consulting fees, lecture fees, travel support, stock or stock options). **Role of the funder:** the manufacturer had no role in the design, conduct, analysis or reporting of the study. |

#### References (supplemental)

*Per the journal's supplemental-data guidelines, references cited in the supplement are listed separately with their own numbering starting at 1, independent of the main reference list.*


1. Bossuyt PM, Reitsma JB, Bruns DE, et al. STARD 2015: an updated list of essential items for reporting diagnostic accuracy studies. BMJ. 2015;351:h5527. doi:10.1136/bmj.h5527
